# ‘Pushing’ versus ‘holding’ isometric muscle actions; what we know and where to go: A scoping and systematic review with meta-analyses

**DOI:** 10.1101/2024.11.04.24316609

**Authors:** Dustin J Oranchuk, André R Nelson, Danny Lum, Alex O Natera, Frank N Bittmann, Laura V Schaefer

## Abstract

**Purpose:** Pushing/pulling isometric muscle actions (PIMA) are widely used to assess strength, fatigue, and neuromechanical factors, whereas holding isometric muscle actions (HIMA) are less understood. However, the distinction between them lacks consensus, complicating application. This study synthesized research comparing PIMA and HIMA to identify potential applications and research gaps.

**Method:** Following registration (PROSPERO: CRD42024530386), databases were searched for studies comparing PIMA and HIMA. Risk-of-bias (revised Joanna Briggs Institute criteria) and certainty (adapted Grading of Recommendations Assessment, Development and Evaluation) were assessed. Meta-analyses examined time to task failure (TTF), ratings of perceived exertion (RPE), heart rate (HR), and mean arterial pressure (MAP); remaining outcomes were synthesized narratively.

**Results:** Fifty-four studies were reviewed (n=915 participants, 374 outcome comparisons). Meta-analysis revealed longer TTF for PIMA at equal absolute intensity (30 outcomes, n=350; *g*=−0.80, *p*<0.001; appendicular muscles: *g*=−0.89, robust to leave-one-out analyses), but longer for HIMA in two axial-muscle studies (*g*=0.61, *p*=0.009). Differences in HR and MAP were trivial (|*g*|≤0.19, *p*≥0.008); RPE was higher during PIMA only at 50% TTF (*g*=−0.31, *p*=0.008). Narrative synthesis (low/very low certainty) associated PIMA with higher peak torques, discharge rates, and D1 inhibition; HIMA with higher burst rates, heteronymous facilitation, glucose uptake, and faster increases in RPE, HR, MAP, EMG amplitude, and force fluctuations.

**Conclusion:** PIMA permits longer sustained activation in appendicular muscles (moderate certainty). Very-low-certainty evidence suggests more complex control strategies for HIMA, raising hypotheses for injury-prevention strategies and novel diagnostic and rehabilitation approaches; prospective and clinical studies, and longitudinal studies comparing PIMA- and HIMA-based training, are warranted.

## Introduction

Different muscle action types (i.e., concentric, isometric, eccentric) are commonly used in research to examine acute physiological responses. They are also essential in clinical and sports performance settings to elicit specific morphological, neuromuscular, and performance adaptations through tailored movements and workloads [1, 2]. While concentric and eccentric actions are often highlighted for their distinct metabolic costs and adaptation profiles, including neuromuscular cross-education, muscle fascicle length, and region-specific hypertrophy [2], the importance of differentiating between two distinct types of isometric muscle actions has been underemphasized. Foundational studies have long demonstrated that different types of isometric tasks exhibit distinct neurophysiological [3-6] and metabolic [7, 8] characteristics. More recent investigations further highlight the need to recognize and distinguish the two types of isometric muscle actions, as they exhibit unique acute responses across multiple neurological and neuromuscular parameters [9-12], potentially influencing outcomes and application [1, 13].

Research examining muscle action types has existed since at least 1895, with Adolph Fick examining cardiac muscle under isometric conditions [14]. However, isometrics are commonly thought of as a single category of muscle action. There has been a recent surge in studies comparing ‘two modes’ of isometric muscle actions [9, 10] and hybrid muscle actions such as ‘quasi-isometrics’ [15-20] and ‘Adaptive Force’ [21-29]. The two distinctive forms are known by several names, including pushing/pulling (PIMA) and holding (HIMA) isometric muscle actions [9], force and position tasks [3], and overcoming and yielding isometrics [30]. Regardless of the semantics, PIMA involves exerting force against an immovable object (e.g., a strain-gauge, a wall, or a power rack). Conversely, HIMA is defined as maintaining a set position while resisting an external force (e.g., gravity, inertially loaded weight, cable stack, training partner, examiner) (**Fig. 1**).

**Fig. 1.**
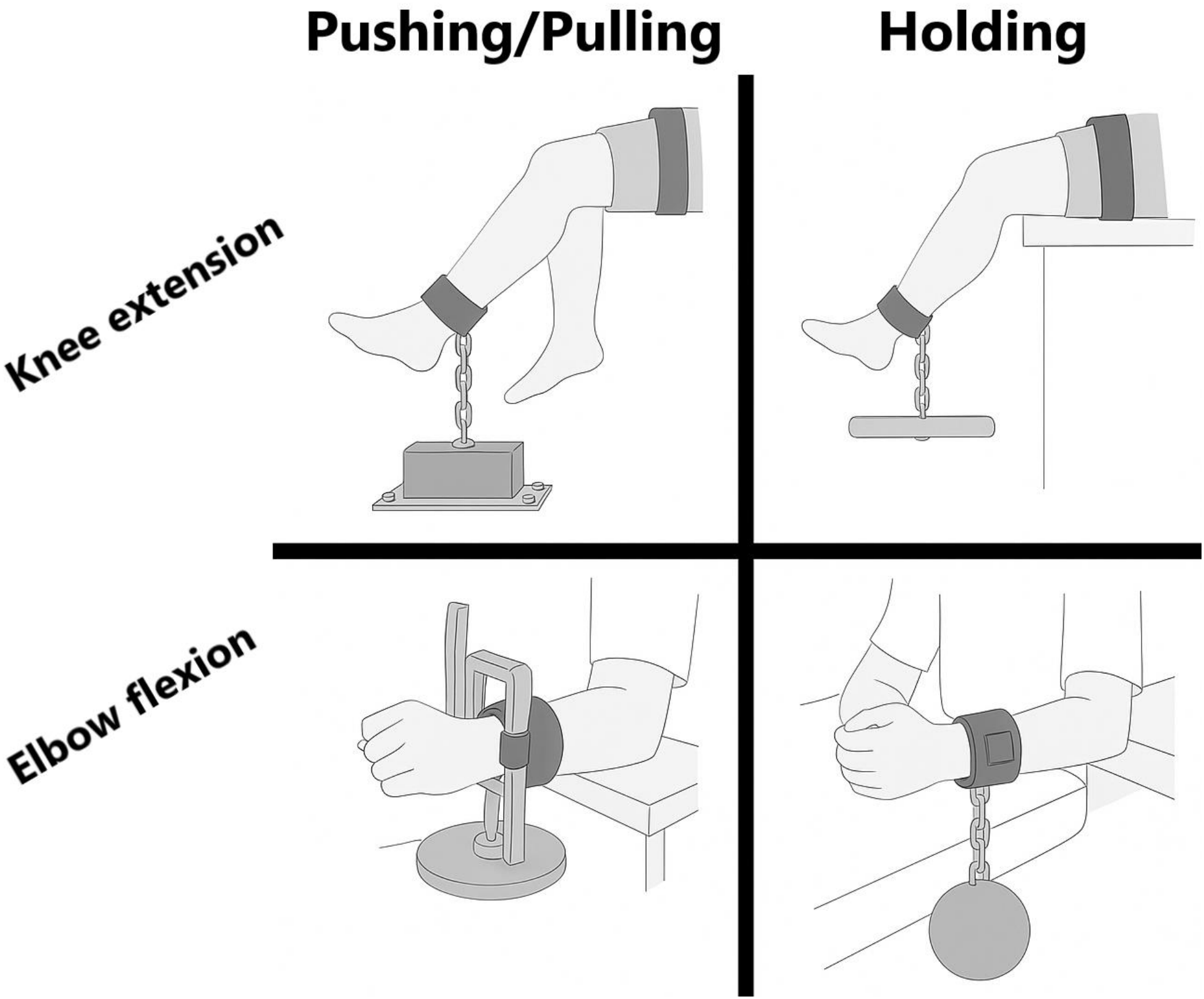
Examples of single-joint pushing/pulling and holding isometric muscle actions. (images adapted from Rudroff et al. [7] and Pascoe et al. [31]).

While PIMAs are far more common in sports medicine and physiological research [1], typically as a means to assess maximal force production capacity [32], several recent studies have utilized HIMA in rehabilitative and performance contexts [33-37]. In parallel with this expanding use, practitioners have shown increasing interest in whether these two forms of isometric muscle action carry distinct practical implications [38-42]. Despite early work demonstrating that force- and position-based tasks differ in neuromechanical control and patterns of fatigue, notable gaps and inconsistencies remain in the literature. Specifically, although multiple studies have reported differences in mechanical [4, 43], neural [9, 11], perceptual [44, 45] and cardiometabolic variables [7, 46, 47], there is still no clear or consistently applied differentiation between PIMA and HIMA, and the physiological, mechanistic and contextual bases for these task-dependent differences remain poorly characterized. Therefore, the primary purpose of this review was to systematically identify, evaluate, and map the existing literature directly comparing PIMA and HIMA, and to quantitatively synthesize outcomes where sufficient comparable data were available, to determine whether objective measures support their distinction. A secondary aim was to use the available evidence to inform recommendations for researchers and practitioners.

## Methods

The protocol for the present review was registered at the National Institute for Health and Care Research (NIHR) PROSPERO (CRD42024530386). This systematic review conformed to the “Preferred Reporting Items for Systematic Reviews and Meta-Analyses” (PRISMA) guidelines [48]. As only published data were analyzed, no Institutional Review Board approval was necessary.

### Inclusion and exclusion criteria

All included studies met the inclusion criteria: 1) published in English; 2) peer-reviewed journal publications or doctoral dissertations; 3) human participants; and 4) evaluated the mechanical, neurological, perceptual, or physiological effects of PIMA and HIMA. Studies were excluded if they: 1) were conference proceedings (papers, posters, presentations); and 2) did not include both PIMA and HIMA.

### Literature search strategy

The initial search utilized CINAHL, Embase, MEDLINE, PubMed and Web of Science databases from inception to January 2024. Terms were searched for within the article title, abstract, and keywords using the Boolean operators ‘OR’ and ‘AND’ with truncation ‘*’. Boolean phrases comprising the search terms were: (isometric* OR static*) AND (contraction* OR action* OR task*) AND (holding OR position) AND (pushing OR force). No limiters or filters were applied. Secondary searches included: a) screening the reference lists of included studies; b) forward citation tracking; c) search alerts to monitor new search results; and d) contacting the most common authors of included outputs. Database searching was re-run regularly, with the final search in August 2025.

### Study selection

Database results were uploaded to the EndNote reference manager (X9.0.3; Clarivate Analytics, Philadelphia, PA) and utilized to remove 3080 duplicates, while 22 duplicates were manually identified and removed. The remaining publications were uploaded to the Rayyan systematic review software, where titles and abstracts were independently screened for eligibility by two authors (DJO, ARN) blinded to each other’s decisions. Three authors (DJO, ARN, LVS) then independently screened the full texts to finalize eligibility for inclusion; studies co-authored by one of them were screened by the other two. All disagreements were resolved through discussion, with the lead author making the final decision.

Because terminology surrounding PIMA and HIMA is inconsistent, all studies were evaluated against the operational definitions established for this review. Specifically, we examined whether the task involved force generation against an external resistance (PIMA) or position maintenance against an externally applied load (HIMA), regardless of the terminology used by the original authors (e.g., “pushing”, “force task”, “holding”, “position task”).

### Data extraction

A spreadsheet was used to record extracted data, including: 1) publication details; 2) participant information; 3) key equipment; 4) condition details; 5) means, standard deviation and, where available, raw data; and 6) reported statistical outputs. Corresponding authors were contacted via email if insufficient data were reported. Four authors (DJO, ARN, DL, LVS) independently completed data extraction. Any differences were resolved via discussion, with the lead author finalizing each decision. Participant numbers refer to participants tested (including those who later dropped out); the number of participants included in the analyses is given as additional information. Where publications reported data from the same cohort, the number of unique participants is additionally given.

WebPlotDigitizer (v4.1; https://automeris.io/WebPlotDigitizer/) was used to extract data from figures when raw values were unavailable. Images were exported from PDFs as high-resolution files. Means were estimated from the midpoint of error bars; when bars were asymmetric (e.g., triangles), the midpoint of the symmetrical error bars (SD or SE) was used instead. Upper error bars were digitized unless obscured by overlapping symbols, in which case lower bars were used. When error bars were embedded within the mean symbol, the largest plausible error was estimated by manually overlaying the bar width, which could overestimate the error for data points with minimal variability. When not otherwise stated, we assumed no missing samples and calculated standard deviations from reported standard errors using the study’s sample size.

### Risk-of-bias and reporting characteristics

Risk of bias was assessed using the 2024 revised Joanna Briggs Institute (JBI) critical appraisal tool for quasi-experimental studies [49]. This tool was selected because the included evidence predominantly comprised acute experimental comparisons, most commonly within-participant PIMA–HIMA designs, rather than longitudinal intervention studies. The nine JBI items were interpreted at the level specified by the revised framework: Q1–Q4 at study level, Q5–Q7 at outcome level, and Q8–Q9 at result level. Full operational definitions and decision rules used are provided in **Online Resource 1**. Limitations concerning task sequence, period and carryover effects are addressed separately.

The original risk-of-bias appraisal was undertaken by four authors (DJO, ARN, DL, AON), with studies allocated so that review authors did not assess primary studies on which they were authors. The nine included studies involving review authors FNB and/or LVS were therefore assessed by non-conflicted reviewers. To align the original appraisal with the 2024 revised JBI framework, Q6 and Q7 were independently reassessed for all 54 studies, while Q3, Q4, Q8 and Q9 were re-examined where the original assessment was incomplete or unclear, or where application of the revised criteria could alter the judgment. Q1, Q2 and Q5 were retained where the original assessment directly addressed the revised JBI item.

For Q6, a definitive “Yes” required the same outcome construct and sufficiently equivalent measurement method, timing, processing and scoring across PIMA and HIMA; task-inherent differences in force-versus position-control procedures were not themselves considered inconsistent measurement. For Q7, standardized procedures, repeated trials, averaging or calibration alone were not considered evidence of reliability; a definitive “Yes” required empirical within-study evidence of reliability or repeatability for the review-relevant outcome(s). Q8 was assessed at the result level and considered whether participant-, limb-, trial-, motor-unit- or outcome-level exclusions or changing denominators could plausibly bias the PIMA–HIMA comparison. Q9 explicitly considered the paired or repeated-measures structure, the true independent sampling unit, clustering of multiple observations within participants, handling of missing or excluded observations, and appropriateness of the statistical model.

Seven studies [3, 9, 50-54] were jointly assessed by DJO and ARN to pilot the appraisal tool and were excluded from the original inter-rater agreement calculation. For the remaining 47 studies, reviewers agreed on 336 of 376 independently double-rated responses across eight appraisal and reporting fields (89.4%; unweighted Gwet’s AC1 = 0.87). Agreement was calculated before adjudication using unweighted AC1 [55]. For the revision recheck, two non-conflicted reviewers (DJO and ARN) independently completed the required assessments, with disagreements adjudicated by a third reviewer (DL) to determine the final judgment. Reviewers agreed on 135 of 150 paired revision assessments (90%; unweighted Gwet’s AC1 = 0.874). The original appraisal coefficient is a descriptive pooled summary across heterogeneous appraisal and reporting fields; the revision calculation included three assessments for which both reviewers recorded “outcome-specific”, treated as a distinct response category.

We summarized participant characteristics and the completeness of demographic and baseline reporting descriptively, separately from the JBI risk-of-bias appraisal, and did not use them to inform JBI judgments or an overall study-quality score. We visualized final JBI judgments as a condensed study display and item-level summary plots.

Certainty was assessed for each quantitatively synthesized outcome and each narratively synthesized outcome group using an adapted GRADE approach [56]. All outcomes entered the assessment at high certainty. This common starting point reflected the predominantly controlled, within-participant comparisons of immediate responses to investigator-imposed PIMA and HIMA conditions: these comparisons reduce confounding by stable individual characteristics and provide a direct task comparison without an independent untreated group. The high-certainty starting point was a review-level adaptation and is not a claim that all task sequences were randomized or that all designs offered equivalent protection against bias. The revised JBI judgments informed the risk-of-bias domain; certainty was downgraded for serious or very serious concerns in risk of bias, inconsistency, indirectness, imprecision, and publication bias. We note that risk-of-bias appraisal using the JBI quasi-experimental checklist does not comprehensively evaluate allocation of task order, period or carryover effects, or selective reporting. Imprecision judgments considered confidence intervals and information size, informed by Core GRADE guidance. We also used a post hoc paired-design information-size benchmark (two-sided α = 0.05, 80% power, *r* = 0.50 and target standardized difference 0.20), with sensitivity to these assumptions. [57]. This descriptive threshold was not a validated clinical minimum important difference; calculations and outcome-specific rationales are reported in **Online Resource 2**. One reviewer (ARN) prepared the GRADE assessments, which were reviewed by a second reviewer (DJO); final ratings and supporting rationales were agreed through discussion. For outcomes without a pooled estimate, we followed the guidance of Murad et al. [58], with synthesis reported using SWiM [59]. Outcome-level ratings and reasons for downgrading or not downgrading are provided in **Online Resource 2**.

### Scoping component of the review

In addition to the systematic review and meta-analyses, this review incorporated a scoping component to comprehensively map the breadth of research comparing PIMA and HIMA. Specifically, we summarized the range of study populations, muscle groups, loading intensities, outcome measures, and experimental methodologies, and identified knowledge gaps in areas where quantitative synthesis was not feasible due to methodological or outcome heterogeneity. Thus, the systematic review addressed outcomes that were synthesized quantitatively (meta-analysis) or, where pooling was not feasible, narratively using structured outcome counts (see Narrative synthesis), whereas the scoping component provided an overview of the broader evidence base and informed recommendations for future research.

### Statistical analysis

#### Calculation of effect sizes

Raw data, effect sizes, and *p*-values were extracted to a spreadsheet. Extracted raw data were used to calculate standardized mean differences (Cohen’s effect size [*d*]) between PIMA and HIMA conditions. Standardized mean differences |*d*| were interpreted as: <0.20 trivial, 0.20–0.49 small, 0.50–0.79 moderate, and ≥0.80 large [60]. Studies already reporting effect sizes were double-checked where possible. For the meta-analyses, *d* was converted to Hedges’ *g* (see Meta-analytic synthesis). Effect sizes and *p*-values are both reported in the supplemental tables for the respective parameters (**Online Resources 3–5)**. Standardized mean differences were calculated for all possible time points and variables using the following equation:

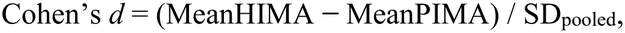

where SD_pooled_ is the square root of the mean of the two condition variances. Positive values indicate higher, negative values lower values during HIMA compared with PIMA. In the narrative synthesis, effect sizes are signed; in the supplemental tables, they are reported as absolute values, with the direction of the difference indicated separately.

Percentage differences between PIMA and HIMA conditions were also calculated and reported where appropriate using the following equation:

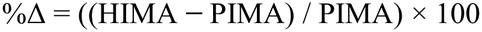

#### Meta-analytic synthesis

Time to task failure (TTF), ratings of perceived exertion (RPE), heart rate (HR) and mean arterial pressure (MAP) were meta-analyzed where outcome definitions, task conditions and available summary data permitted comparable estimates. Other outcomes were synthesized narratively because measures, experimental protocols or reporting were insufficiently comparable for the intended synthesis, often with few studies per parameter.

Meta-analytical procedures (effect sizes, within-participant variances, random-effects models, meta-regression, and sensitivity analyses) were implemented in Microsoft Excel 365 (Microsoft Corporation, Redmond, WA, USA); the pooled estimates and sensitivity analyses were additionally computed in Python (Version 3.13.16; NumPy 2.5.3, SciPy 1.18.1) and reproduced the spreadsheet results.

Each study examining TTF provided the data required to calculate standardized mean differences between PIMA and HIMA (means and standard deviations or standard errors). RPE, HR, and MAP data were often reported in figures and extracted (see data extraction). Data were transferred directly to the calculation workbooks to avoid potential human error, and standardized mean differences were confirmed by manual calculation. Hedges’ *g* (Cohen’s *d* corrected for small-sample bias) was calculated as HIMA − PIMA; negative values indicate lower values during HIMA, and the forest plots show signed values. Because the comparisons included in these meta-analyses used within-participant designs, the sampling variance of *g* accounted for the within-participant correlation *r*: Var(g) = 2(1 − *r*)/n + g^2^/[2(2n − 3.94)]. None of the primary studies reported *r*. For TTF, *r* was back-calculated from the reported test statistics where possible (smallest compatible *r*, but not less than 0.5, where only a threshold *p*-value was given) and otherwise set to 0.5; for RPE, HR and MAP, *r* = 0.5 was used for all outcomes. The influence of *r* was examined in sensitivity analyses (**Online Resource 6**). In the variance formula, n is the number of participants contributing to that paired comparison. Summary participant totals count participants once within each study.

Including multiple effects from a single study violates the assumption of independence, as effects from the same study are likely to be more similar than those from other studies and would influence statistical power. However, when only a small proportion of studies report multiple outcomes, dependence might be ignored [61, 62]. For TTF, four of 23 studies reported multiple outcomes with overlapping participants at different intensities [8, 63], forearm positions [64], or muscle groups [53]. The meta-analysis was therefore computed with both single and combined outcomes to examine whether this dependence affected the pooled estimate. The forest plots illustrate single outcomes, while statistical results are provided for both. For the combined analysis, the effect sizes of a single study obtained at multiple loading intensities or forearm orientations were averaged as a single comparison [53]; outcomes from different muscle groups of the same study were kept separate because they belong to different subgroups.

A random-effects model with restricted maximum likelihood (REML) estimation of the between-study variance was chosen because of substantial inter-study variability in age, sex, muscle groups, and loading intensities. Effect sizes are reported as Hedges’ g with 95% confidence intervals in square brackets and were interpreted based on their absolute value |g|: <0.20 trivial, 0.20–0.49 small, 0.50–0.79 moderate, and ≥0.80 large [60]. Exploratory meta-regression (bubble plots) and subgroup analyses (Q test for between-subgroup differences) were employed to examine the effect of loading intensity and muscle group; RPE, HR, and MAP were analyzed separately at task initiation, 50% of TTF, and task failure. Muscle groups were ordered by typical muscle mass and coded: 1-finger abductors, 2-wrist extensors, 3-ankle dorsi-flexors, 4-elbow flexors, 5-elbow extensors, 6-knee extensors, and 7-trunk extensors [65, 66].

Robustness of the pooled estimates was examined by sensitivity analyses for TTF, the outcome with the largest evidence base, in the appendicular subgroup (28 outcomes from 22 studies); results for all outcomes are provided in **Online Resource 7**. First, we performed leave-one-out analyses at the level of individual publications and first authors. Second, because a substantial proportion of outcomes originate from a small number of research groups, we performed leave-one-laboratory-out analyses, in which we removed the outcomes of each laboratory in turn. We defined laboratories as the institution with which all or most authors of a publication were affiliated, as a proxy for the site of data collection; we also jointly removed the two laboratories that collaborated directly on included studies (Colorado Boulder and Brussels). The pooled estimate and τ^2^ were re-estimated by REML in each analysis.

#### Statistical heterogeneity

Between-study heterogeneity was quantified using the between-study variance τ^2^, Cochran’s Q test, and the I^2^ statistic, which represents the percentage of total variation in effect estimates attributable to heterogeneity rather than sampling error; I^2^ was derived from τ^2^ and the typical within-study sampling variance. I^2^ values were interpreted as: <50% low, 50–74% moderate, and ≥75% high heterogeneity [67]. For TTF, the funnel plot was visually examined for asymmetry or multiple outcomes outside the funnel, which could indicate small-study effects or publication bias [68].

#### Narrative synthesis

Outcomes that were reported in too few studies or with incompatible methodologies to permit meta-analysis were synthesized narratively, following the Synthesis Without Meta-analysis (SWiM) reporting guideline **[59]**. Outcome-level comparisons were grouped by parameter; each comparison within a study (e.g., at different intensities, muscle groups, or age groups) was counted separately (see Summary of isometric muscle action differences). For each parameter, we summarized the direction and consistency of effects together with standardized effect sizes where available. Unless otherwise stated, a comparison was counted as favoring PIMA or HIMA when |*d*| or |*g*| ≥ 0.20, irrespective of statistical significance, and as trivial when <0.20. Statistical significance, as given in the primary studies, is reported separately and was not used to determine the direction of effects, unless no effect size could be derived. We described parameters examined in only one study individually. Counts of comparisons favoring PIMA, HIMA, or showing trivial effects describe the consistency of the available evidence; they do not provide a quantitative estimate of effect or statistical support and should be interpreted as structured narrative evidence rather than pooled estimates.

## Results

The systematic literature search (**Fig. 2**) yielded 54 studies fulfilling the inclusion criteria [3-12, 21-26, 31, 43-47, 50-54, 63, 64, 69-93]. Forty-two studies were found through database searching [5, 6, 8-12, 21-26, 31, 44-47, 50, 53, 54, 63, 70, 71, 73-89, 93], while 11 publications were included following reference searches [3, 7, 43, 51, 52, 64, 69, 72, 90-92]. Since nine of these were from the same laboratory group [7, 43, 52, 64, 69, 72, 90-92], the primary investigator of the relevant laboratory was contacted, resulting in an additional study [4].

**Fig. 2.**
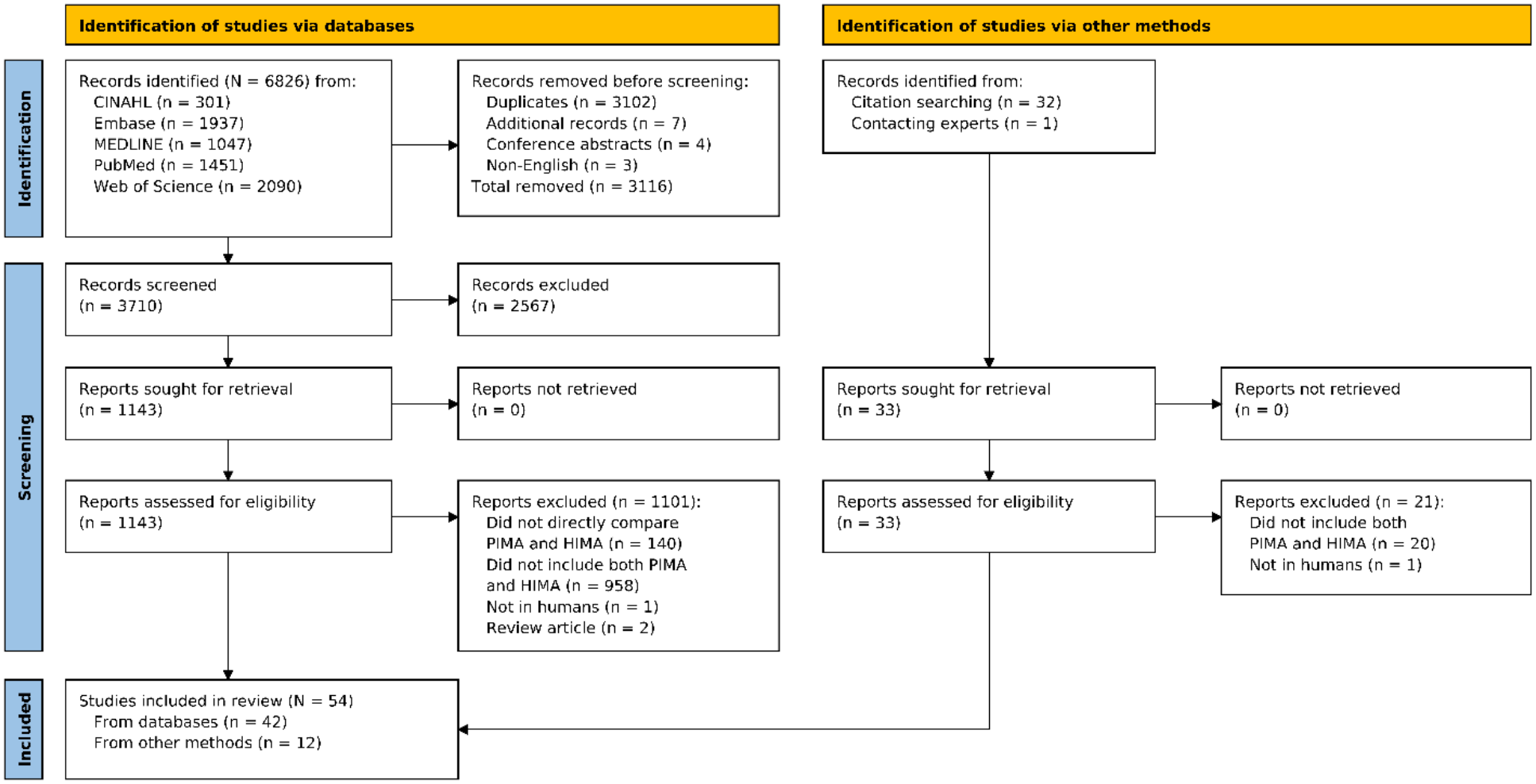
PRISMA diagram summarizing the literature search strategies.

### Study characteristics

The 54 studies comprised 915 tested participants (619 male, 263 female, sex not reported for 33; mean age of study samples 29.6 ± 10.5 years) [3-12, 21-26, 31, 43-47, 50-54, 63, 64, 69-93]. Of these, 892 participants were included in the analyses. Because three pairs of publications reported data from the same cohorts [6, 45, 80, 81, 87, 88], the number of unique participants was ∼871 (578 male, 260 female). The publication years ranged from 1995 [50] to 2024 [21] with mean publication year of 2013 ± 6.9. Demographic data are approximate because sex [50, 69, 85] and age [50, 69] were missing or only partially reported in several studies. Ten studies were exclusively male [5-7, 12, 21-23, 26, 64, 87], 41 were mixed sex [3, 4, 8-11, 24, 25, 31, 43-47, 51-54, 63, 70-84, 86, 88-93], with three not reporting sex [50, 69, 85]. Forty-six studies examined young adults (mean: 21–33.5 years) [3-6, 8-12, 22-26, 43-47, 51-54, 63, 64, 70, 72-76, 79-93], two included older participants (mean: 64–72 years) [21, 78], four young and old sub-groups (mean: 47–52 years) [7, 31, 71, 77], with two not reporting age [50, 69]. Single studies concentrated on specific samples, such as strength and endurance athletes [22], soccer players [23], patients with knee osteoarthritis [21], or participants undergoing experimental joint pain [44, 45].

All 54 studies utilized single-joint actions at the same absolute load [3-12, 21-26, 31, 43-47, 50-54, 63, 64, 69-93]. The most common agonist muscle group was the elbow flexors (n = 24) [3, 4, 6, 8, 22-24, 31, 47, 50, 53, 54, 63, 64, 72, 75, 76, 78, 80, 81, 83, 87, 90, 92], followed by the knee extensors (n = 8) [7, 12, 44, 45, 73, 88, 89, 91]. Finger abductors (n = 6) [5, 11, 52, 70, 82, 85], wrist extensors (n = 3) [43, 69, 71] and flexors (n = 2) [46, 69], ankle dorsi-(n = 3) [77, 79, 86] and plantar-flexors (n = 3) [51, 74, 84], elbow extensors (n = 5) [9, 10, 25, 26, 50], hip/trunk extensors (n = 2) [53, 93], knee flexors (n = 1) [21], and hip flexors (n = 1) [23] were also agonist muscle groups investigated. Bittmann et al. [24] (elbow/hip flexion), Thomas et al. [53] (trunk extension/elbow flexion) and Buchanan and Lloyd (elbow flexion/extension) [50] compared PIMA and HIMA across multiple agonist groups. One study included different forearm postures during elbow flexions [64], while another examined elbow flexion with supinated and neutral forearms [64, 90]. Muscle action intensities differed widely, ranging from 3.8–100% (mean: ∼34.6 ± 27.8%) of pushing or pulling MVIC. Most studies (n = 40) investigated intensities ≤30% MVIC [3-8, 11, 31, 43-45, 50-54, 63, 64, 69-79, 82-85, 87-93], while 14 studies investigated intensities of 40–90% MVIC [5, 8-10, 12, 26, 46, 47, 63, 70, 74, 80, 81, 86], and five studies used adaptive muscle action of >100% MVIC (gradual increase in applied force from submaximal to supramaximal intensities in one trial) [21-25].

Twelve included multiple intensities [3, 5, 8, 10, 31, 50, 63, 70, 71, 74, 80, 84]. Isometric tasks were taken to failure in 30 studies [4, 5, 7-10, 21-26, 43, 47, 53, 54, 63, 64, 72, 73, 75, 77-79, 81, 83, 89, 91-93].

### Risk of bias and reporting characteristics

The revised JBI appraisal is presented item-by-item in **Fig. 3** and **Online Resource 1**. The basic PIMA–HIMA comparison was generally well defined across the included literature, while the most frequent source of uncertainty concerned whether reliability of the review-relevant outcome measures had been demonstrated empirically within the study. Many reports described standardized acquisition procedures, repeated trials, averaging or calibration but did not provide within-study reliability or repeatability evidence sufficient for a definitive “Yes” judgment for JBI Q7.

**Fig. 3.**
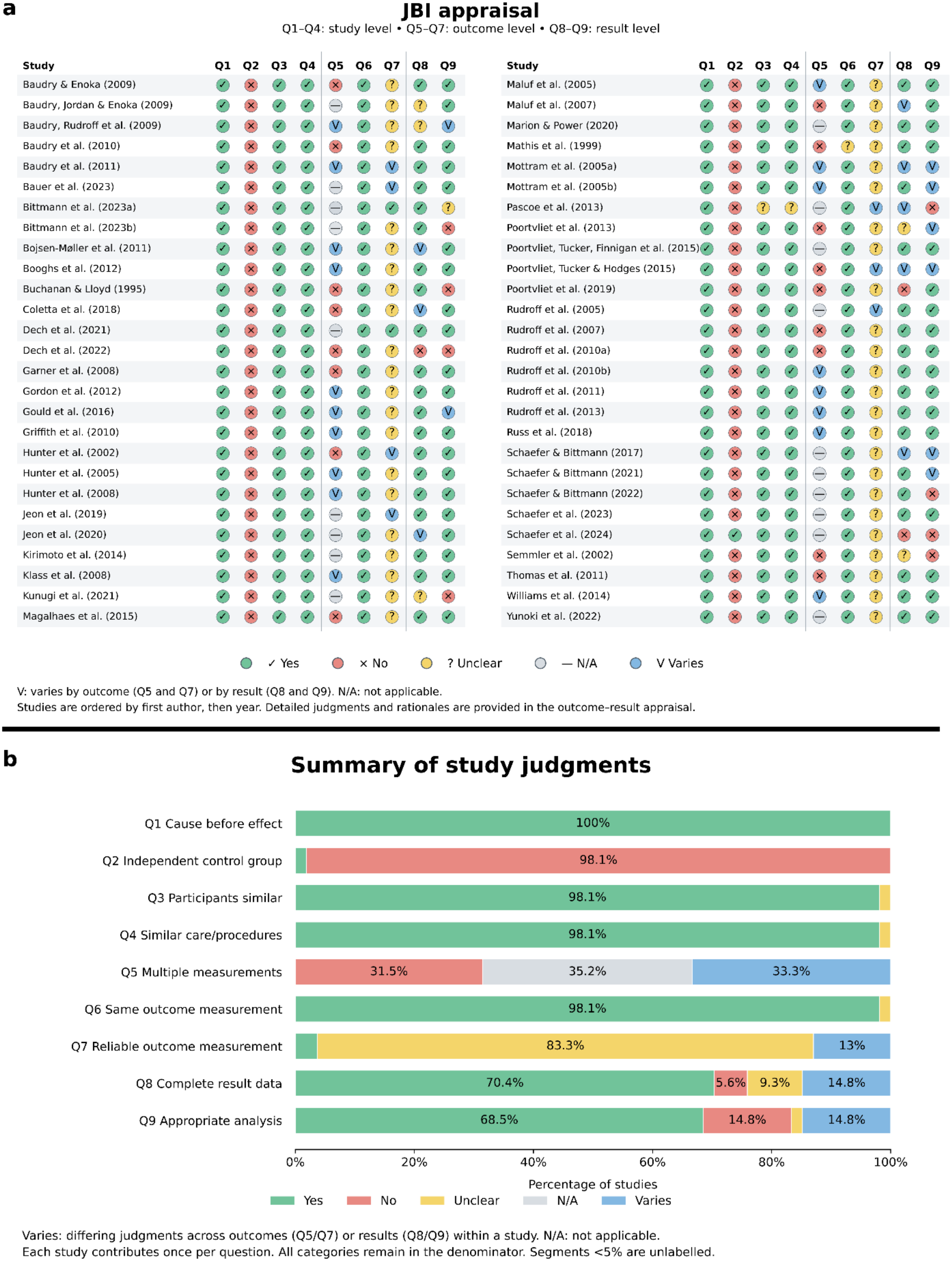
Risk-of-bias appraisal of the 54 included studies using the revised JBI critical appraisal tool. **(a)** Condensed study-level display of judgments for study-level items (Q1–Q4), outcome-level items (Q5–Q7), and result-level items (Q8–Q9). Yes, No, Unclear, and N/A (not applicable) are formal judgments. V (Varies) indicates differing judgments across outcomes (Q5–Q7) or results (Q8–Q9) within a study and is a display category rather than an additional JBI response category. **(b)** Study judgments are summarized as percentages, with each study contributing once per question (denominator = 54 for every item). All displayed categories, including N/A and Varies, remain in the denominator. Detailed judgments and rationales are provided in Online Resource 1. Segments representing <5% are unlabeled.

Participant reporting also varied considerably across studies. Five studies lacked detail regarding study participants [3, 50, 51, 69, 74], and were substantially older (2005.2 ± 8.1, 1995–2015) than the overall mean publication year (2013 ± 6.9), suggesting improved reporting in more recent research. Neither Buchanan and Lloyd [50] nor Baudry and Enoka [69] reported sex, age, height, mass or other demographic or baseline characteristics. Inclusion and exclusion criteria, physical activity level, and baseline characteristics (e.g., strength, power) were not reported by Garner et al. [74]. Neither Magalhães et al. [51] nor Mathis et al. [3] reported height, mass, inclusion/exclusion criteria, or any baseline characteristics.

### Certainty of evidence

Under the adapted GRADE approach, certainty was moderate for appendicular TTF, HR and MAP; low for overall and axial TTF, RPE at midpoint, rates of increase of RPE/HR/MAP, and force/position fluctuations; and very low for maximal torque/MVIC decline, and the grouped neurophysiological/neuromuscular evidence. Full domain judgments and supporting rationales are presented in **Online Resource 2**.

### Summary of isometric muscle action differences

A descriptive overview of the primary outcomes, including the total number and parameters of comparisons (e.g., TTF, RPE increases, discharge rates) and the frequency of statistically significant or potentially meaningful differences, is presented in **Table 1**. The sustained-task comparisons generally matched absolute target load and joint position between PIMA and HIMA; maximal-capacity and other distinct paradigms are described separately. In total, 374 outcome comparisons were identified. Where statistical significance was reported, 59 outcomes were higher for PIMA (*p* < 0.05), 98 were higher for HIMA, and 202 were not statistically significant. When effect sizes were considered (where available) irrespective of *p*-values, the balance shifted: 86 comparisons favored PIMA, 86 favored HIMA, and 42 were trivial (|*d*| or |*g*| < 0.20). This narrowing partly reflects that several outcomes significantly higher during HIMA lacked the data to compute an effect size, while a number of statistically non-significant comparisons nonetheless showed non-trivial effects, several favoring PIMA.

**Table 1.** Structured narrative summary across synthesized outcomes, reporting outcome-level results of the individual studies (number of outcomes, direction of statistically significant differences (*p* < 0.05) and standardized effect size)

|  | **Number of outcomes** |  |  |  |  |  |  |
|---|---|---|---|---|---|---|---|
|  | **Total** | **Significant *p*-value** |  |  | **Effect size** |  |  |
| **Parameter** |  | HIMA < PIMA | HIMA > PIMA | n.s. | HIMA < PIMA | HIMA > PIMA | trivial effect |
| TTF<sup>†</sup> | 30 | 21 | 3 | 6 | 24 | 3 | 3 |
| Fluctuations | 14 | 7 | 3 | 4 | 10 | 3 | - |
| Increase fluctuations | 23 | 5 | 13 | 5 | 9 | 14 | 0 |
| Peak torque | 3 | 3 | 0 | 0 | 3 | 0 | - |
| MVIC decline* | 28 | 0 | 3 | 25 | 7 | 7 | 9 |
| RPE 50% TTF<sup>†</sup> | 14 | - | - | - | 8 | 3 | 3 |
| RPE 100% TTF<sup>†</sup> | 15 | 0 | 0 | 15 | 2 | 2 | 11 |
| RPE increase | 16 | 0 | 9 | 6 | - | - | - |
| aEMG agonists | 48 | 5 | 8 | 35 | 4 | 3 | - |
| aEMG synergists | 15 | 2 | 2 | 11 | 1 | 2 | - |
| aEMG antagonists | 17 | 1 | 0 | 16 | - | - | - |
| Increase aEMG agonists | 17 | 0 | 8 | 9 | - | 6 | - |
| Increase aEMG synergist | 10 | 0 | 5 | 5 | - | 2 | - |
| Increase aEMG antagonists | 9 | 0 | 4 | 5 | - | 3 | - |
| Coactivation | 12 | 0 | 1 | 11 | - | - | 1 |
| Coactivation increase | 2 | 0 | 2 | 0 | - | - | - |
| EMG burst rate | 4 | 0 | 2 | 2 | 0 | 2 | 2 |
| Burst rate increase | 3 | 0 | 3 | 0 | 0 | 2 | 0 |
| Discharge rate | 12 | 8 | 0 | 4 | 7 | 0 | 0 |
| CV of ISI | 4 | 1 | 2 | 1 | 1 | 2 | - |
| CV of ISI increase | 3 | 0 | 3 | 0 | - | 2 | - |
| D1 inhibition | 7 | 5 | 1 | 1 | 5 | 1 | - |
| Heteronymous reflex | 12 | 1 | 8 | 3 | - | 7 | 1 |
| HR at end<sup>†</sup> | 14 | 0 | 0 | 14 | 5 | 3 | 6 |
| HR increase | 14 | 0 | 8 | 6 | - | 6 | - |
| MAP at end<sup>†</sup> | 14 | 0 | 1 | 13 | 0 | 8 | 6 |
| MAP increase | 14 | 0 | 9 | 5 | - | 5 | - |
| **TOTAL** | **374** | **59** | **98** | **202** | **86** | **86** | **42** |

Counts represent outcome-level results of the individual studies and are not meta-analytic estimates. Statistical significance is as reported in the primary studies; effect sizes were either taken from the primary studies or, where not reported, calculated from the reported means and standard deviations. Because most primary studies used paired comparisons, the significance counts may differ from the *p*-values shown in the forest plots. Parameters with too few comparable findings, or studies with distinct experimental paradigms, are described narratively in the text and are not included here.
Effect sizes were classified as relevant when Cohen's |*d*| or Hedges' |*g*| ≥ 0.20 and as trivial when < 0.20.
<sup>†</sup> also meta-analyzed, see Figs. 4, 5, 7 and 8; counts refer to the results reported in the individual studies, not to the pooled estimates.
\*Irrespective of epoch of TTF (90% or 100% TTF)
In case the sum of results is not equal to the total number of outcomes the information was not available.
Abbreviations (alphabetical): aEMG = EMG amplitude, CV = coefficient of variation, EMG = electromyography, HIMA = holding isometric muscle action, HR = heart rate, ISI = interspike interval, MAP = mean arterial pressure, MVIC = maximal voluntary isometric contraction, n.s. = non-significant, PIMA = pushing isometric muscle action, RPE = rating of perceived exertion (scale 0–10), TTF = time to task failure

### Performance and mechanical parameters

The 35 studies examining performance and biomechanical differences between PIMA and HIMA are detailed in **Online Resource 3** [4-10, 12, 21-25, 43, 45, 47, 53, 54, 63, 64, 72, 73, 75-79, 81, 83, 87, 89-93]. Twenty-three studies compared TTF [4, 5, 7-9, 43, 47, 53, 54, 63, 64, 72, 73, 75, 77-79, 81, 83, 89, 91-93], while one study investigated the failure rate during paired personal interactions [10]. Twenty-two studies quantified force or position fluctuations or variability throughout each condition [4-6, 8, 12, 43, 45, 63, 64, 72, 73, 75-79, 87, 89-93], 20 studies measured MVIC reductions [5-8, 22, 43, 54, 63, 72, 73, 75-79, 83, 87, 90, 91, 93], five compared the maximal muscle action force capacity [21-25] and 13 compared RPE [4, 5, 54, 63, 64, 73, 75-79, 87, 91].

### Time to task failure

Twenty-three studies (30 outcomes, n = 350) investigating TTF showed almost exclusively longer TTFs for PIMA (718 ± 602 s) vs. HIMA (589 ± 720 s), with a mean percentage difference of −22.5 ± 30.1% (range: −56.2 to 61.5%) [4, 5, 7-9, 43, 47, 53, 54, 63, 64, 72, 73, 75, 77-79, 81, 83, 89, 91-93]. Although single outcomes were based on partially overlapping or similar participants, the methods (high/low intensities, different forearm positions, different muscles) and results differed vastly. Thus, a combination of outcomes would result in considerable information loss, including leveling of single outcomes. According to Becker [62] and Park and Beretvas [61], dependence can be ignored if only a few studies report multiple outcomes. Comparing the meta-analysis with single vs. combined outcomes revealed only minor differences (*g* = −0.80 vs. −0.73), indicating that the dependence between multiple outcomes from the same study did not materially affect the pooled estimate. To avoid loss of information, the forest plot (**Fig. 4a**) illustrates the 30 single outcomes across 23 studies. TTF was significantly longer during PIMA with a moderate to large effect (*g =* −0.80 [−1.11, −0.49], *p* < 0.001). This significant effect is highlighted by all but two appendicular-focused outputs falling to the left of the neutral (grey) line, the two exceptions being elbow flexor outcomes [54, 63]. Combining outcomes provided a similar result (*g* = −0.73 [−1.03, −0.43], *p* < 0.001; 25 outcomes, n = 350).

**Fig. 4.**
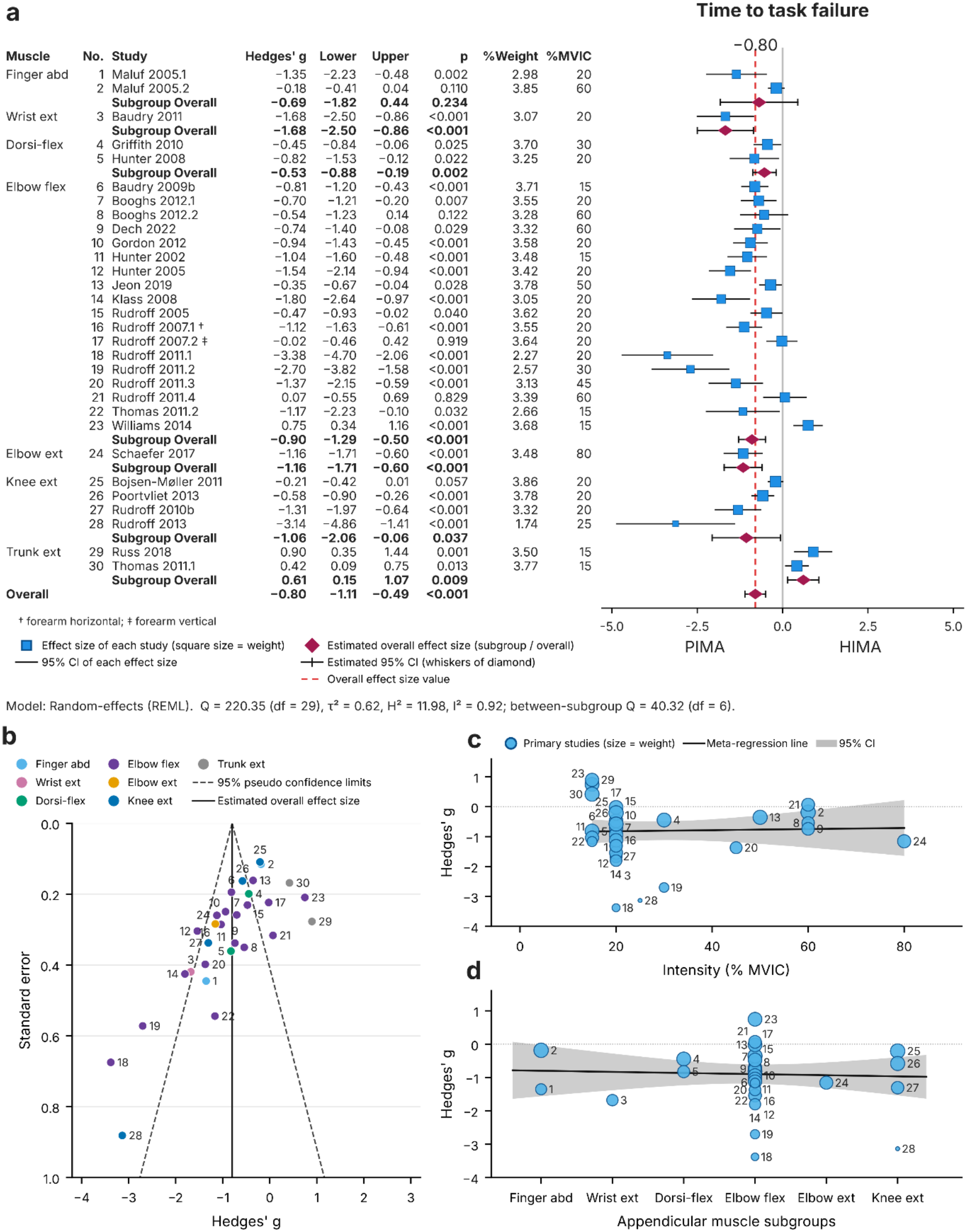
Meta-analysis of time to task failure (TTF) in PIMA vs. HIMA at matched absolute target forces. (**a**) Forest plot of Hedges’ g (negative values: shorter TTF during HIMA); random-effects model with paired variance. (**b**) Funnel plot. Bubble chart regressions for (**c**) loading intensity and (**d**) appendicular muscle subgroups. *p*-values refer to the standardized effect sizes.

Twenty-six of 30 outcomes demonstrated shorter TTF for HIMA with percentage differences from −56.2% to −1.3% (*g* = −3.38 to −0.02). Twenty-four of these showed relevant effect sizes (|*g*| ≥ 0.20) [4, 5, 7-9, 43, 47, 53, 63, 64, 72, 73, 75, 77-79, 81, 83, 89, 91, 92], while two were trivial (|*g*| < 0.20) [5, 64]. The latter occurred for the first dorsal interosseous with intensities of 60% MVIC [5] and elbow flexors with vertical forearm position at 20% MVIC [64]. The remaining four outcomes showed the opposite (longer TTF for HIMA) with percentage differences of 1–61% (*g* = 0.07–0.90) [53, 54, 63, 93]. Two of these studies examined axial muscles (trunk extensors) and showed longer TTF for HIMA with a moderate pooled effect (*g* = 0.61 [0.15, 1.07], *p* = 0.009; 2 outcomes, n = 34) [53, 93]; effects differed significantly between the seven muscle-group subgroups (Q = 40.3, df = 6, *p* < 0.001).

For appendicular muscles, 26 of 28 outcomes showed shorter TTF for HIMA (24 with non-trivial effect sizes); only Williams et al. [54] reported a relevant opposite effect (54%, *g* = 0.75, 15% MVIC, elbow flexors), and Rudroff et al. [63] found similar TTFs at 60% MVIC (%Δ = 1%, *g* = 0.07, elbow flexors). Meta-analysis of TTF with only appendicular muscles, including all outcomes, demonstrated a large effect favoring PIMA (*g* = −0.89 [−1.17, −0.61], *p* < 0.001; 28 outcomes, n = 316). However, in the three studies that tested multiple intensities, the PIMA–HIMA difference in TTF progressively narrowed as absolute load increased [5, 8, 63]. Across the three studies assessing multiple intensities, non-weighted Hedges’ *g* values were large and negative at 20% MVIC (−3.38, −1.35, −0.70) but contracted markedly at 60% MVIC (0.07, −0.18, −0.54), indicating attenuation of the PIMA–HIMA difference with increasing load (**Fig. 4a**). However, these trends are contradicted by Schaefer et al. [9], who utilized high intensities (80% MVIC), and reported significantly longer TTF with PIMA vs. HIMA (*g* = −1.16).

The heterogeneity of the above meta-analyses was high for the appendicular, combined, and all-outcome analyses (I^2^ = 89%, 91%, and 92%) and moderate for the axial subgroup (I^2^ = 54%), supporting the use of the random-effects model. Visual inspection of the funnel plot showed no systematic asymmetry among the more precise estimates (**Fig. 4b**). Subgroup analysis and meta-regression bubble plots determined negligible effects of loading intensity (all outcomes; *p* = 0.837, **Fig. 4c**) and appendicular muscle group (*p* = 0.935, **Fig. 4d**). It must be noted that 23 of 30 investigations used intensities ≤30% MVIC [4, 5, 7, 8, 43, 53, 54, 63, 64, 72, 73, 75, 77-79, 83, 89, 91-93]. Of the remaining seven outcomes (45–80% MVIC), five showed small to large effect sizes (*g* = −1.37 to −0.35) [8, 9, 47, 63, 81], while two were trivial [5, 63].

Leave-one-out analyses indicated that the pooled appendicular estimate did not depend on any individual study: removing each publication in turn yielded pooled effects between *g* = −0.77 and −0.93, and removing all outcomes of each first author in turn yielded *g* = −0.74 to −0.93, with *p* < 0.001 throughout (**Online Resource 7**). Sixteen of the 28 appendicular outcomes originated from a single laboratory (Colorado Boulder). Omitting any single laboratory left the effect statistically significant and in the same direction: removing the Colorado Boulder laboratory reduced its magnitude by approximately one third (*g* = −0.58 [−0.92, −0.24], *p* < 0.001; 12 outcomes, n = 147), whereas omitting any other laboratory yielded *g* **=** −0.89 to −0.93. Omitting the two collaborating laboratories jointly (Colorado Boulder and Brussels) gave *g* = −0.46 [−0.84, −0.09], *p* = 0.016 (9 outcomes, n = 121); heterogeneity remained similar to the full set (I^2^ = 85–86% vs. 89%). Because the Colorado Boulder laboratory contributed 13 of the 21 outcomes at ≤30% MVIC, laboratory and loading intensity could not be separated. The effect was larger at ≤30% MVIC (*g* = −1.02 [−1.39, −0.66]; 21 outcomes) than at ≥50% MVIC (*g* = −0.45 [−0.78, −0.12]; 6 outcomes), although the meta-regression showed no linear association; omitting the Colorado Boulder laboratory from the low-intensity outcomes reduced the estimate to a similar magnitude (*g* = −0.55 [−1.05, −0.06], *p* = 0.029; 8 outcomes; **Online Resource 7**).

Related to TTF, Schaefer and Bittmann examined the ‘failure rate’ during personal interaction of the elbow extensors at 90% MVIC (one partner performed PIMA and the other performed HIMA, in randomized order) [10]. In most trials (85%, *p* < 0.001), the partner performing HIMA left the isometric position first, indicating that the pushing partner could maintain the set position longer, supporting the above results for higher intensities.

### Fluctuations of force and position

Twenty-two studies (37 outcomes) investigated fluctuations of force or position [4-6, 12, 63, 64, 72, 73, 75-78, 87, 89-93]; the outcomes of one study [45] (absolute SD of force vs. position) were not comparable between tasks and are not counted. Eight studies (14 outcomes, Table 1) examined fluctuations (CV or SD of force) at defined time points or averaged over the entire trial [5, 8, 12, 43, 45, 63, 76, 78, 79, 89]. Of these, 10 outcomes showed relevant lower fluctuations in HIMA (*d* = −2.76 to −0.62) [5, 63, 75, 76], three showed higher fluctuations in HIMA (*d* = 0.91–4.05) [12, 78], and one was stated as non-significant with no effect size reported (20% MVIC) [89].

Fifteen studies (23 outcomes, Table 1) compared fluctuation increases between muscle actions (relative to the initial value or as rate of increase) [4, 6, 8, 43, 64, 72, 73, 75, 77, 79, 87, 90-93]. Of these, 14 outcomes showed relevant higher increases for HIMA (*d* = 0.36–2.88) [4, 6, 64, 72, 75, 77, 79, 87, 90, 92], and nine showed lower increases for HIMA (*d* = −1.40 to −0.22) [8, 43, 73, 77, 90, 91, 93].

While difficult to quantify without meta-analysis, absolute fluctuations (CV) were mostly lower in HIMA (10 vs. 3 outcomes), whereas the relative increase in fluctuations over time was more often higher in HIMA (14 vs. 9 outcomes). In the studies that compared several target forces (20–60% MVIC), the direction of the between-task difference was the same at all intensities (lower for HIMA) [5, 8, 63].

### Maximal torques and strength declines

Only one research group compared the maximal torques of HIMA (HIMA_max_) and PIMA (PIMA_max_ = MVIC) [21-25]. Three studies showed significantly lower HIMA_max_ in healthy participants for elbow extensors (*d* = −0.74) [25], elbow flexors (*d* = −2.37) [22], and knee flexors (*d* = −2.62) [21]. Of 20 studies measuring MVIC reductions, 19 studies (28 outcomes) investigated MVIC declines pre-post HIMA or PIMA tasks to failure [5-8, 43, 54, 63, 72, 73, 75-79, 83, 87, 90, 91, 93] with inconsistent results. Seven outcomes revealed a larger MVIC decline for HIMA (*d* = 0.23 to 2.6, 9.6–50.0%) [7, 8, 63, 72, 90, 93] and seven a smaller decline for HIMA (*d* = −0.73 to −0.2, −40.2 to −17.2%) [6, 8, 63, 75, 77, 87]. Only three outcomes reached significance, all showing larger MVIC declines for HIMA after 90% of TTF [7, 72]. Separately, one study examined the decline in maximal torque (PIMA_max_ vs. HIMA_max_) after 30 repeated adaptive supramaximal HIMA trials, with HIMA showing a 133% greater decline than PIMA [22].

### Ratings of perceived exertion

Eleven studies investigated RPE during TTF trials [4, 5, 54, 63, 64, 73, 75, 77-79, 91], whereas two further studies reported RPE throughout a trial not taken to failure and were therefore not included in meta-analysis [76, 87]. Meta-analyses of 14–16 outcomes (9–11 studies, 27.7 ± 15.1% MVIC, range 15–60%) found no significant difference in RPE at initiation (n = 196, *g* = 0.05 [−0.10, +0.19], *p* = 0.544) [4, 5, 54, 63, 64, 73, 75, 77-79, 91] or at task failure (n = 176, *g* = 0.01 [−0.13, +0.14], *p* = 0.941; **Fig. 5a**) [4, 5, 54, 63, 64, 73, 77-79, 91]. At 50% TTF, however, RPE was significantly lower during HIMA with a small effect (n = 166, *g =* −0.31 [−0.54, −0.08], *p* = 0.008) [4, 5, 63, 64, 73, 77-79, 91]. This result was robust to the assumed within-participant correlation (*r* = 0, 0.3, 0.5, 0.7; *p* = 0.007–0.010; **Online Resource 6**)

**Fig. 5.**
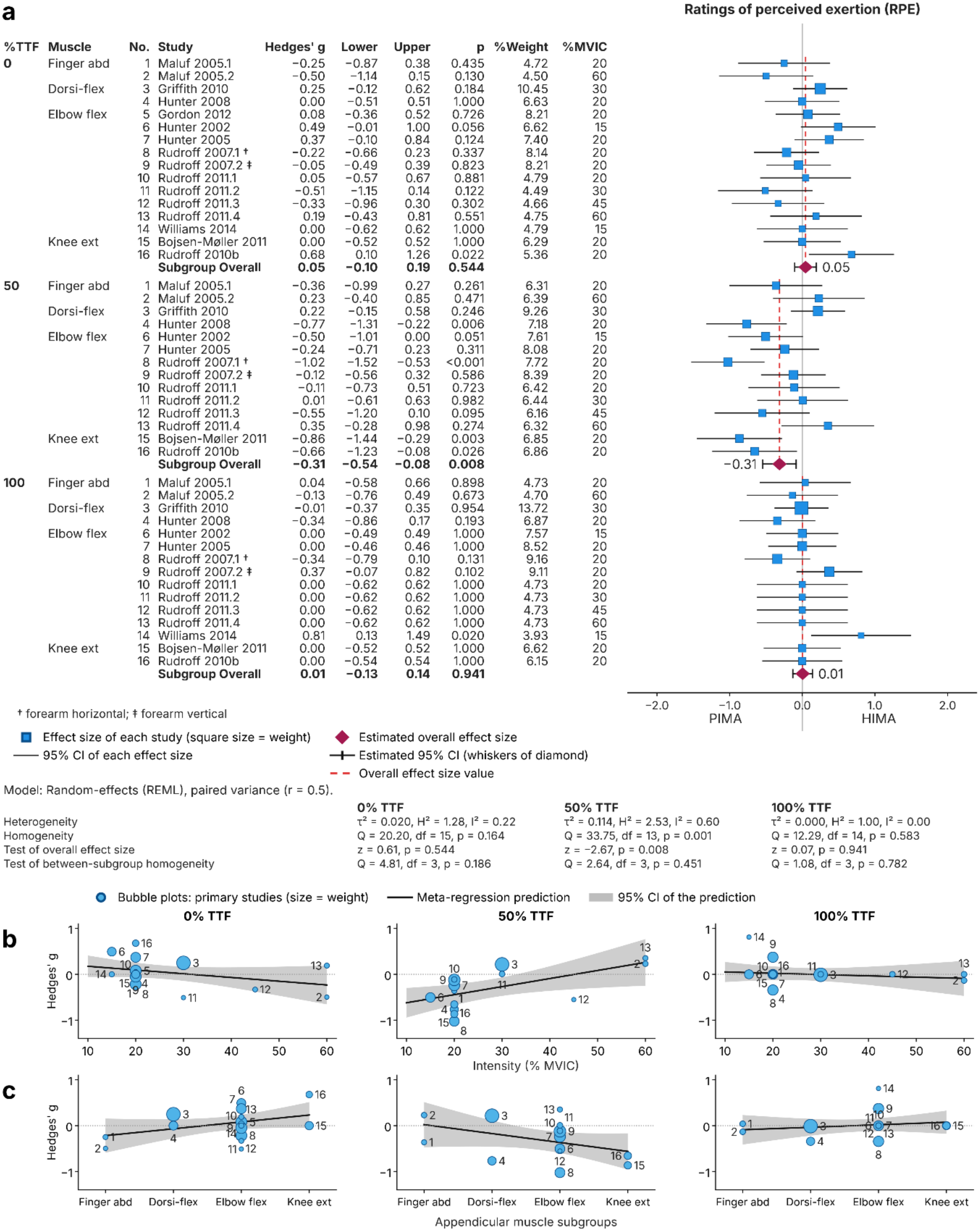
Meta-analysis of ratings of perceived exertion (RPE) in PIMA vs. HIMA at 0%, 50% and 100% of time to task failure (TTF). (**a**) Forest plot of Hedges’ g (negative values: lower RPE during HIMA); random-effects model with paired variance. Bubble chart regressions for (**b**) loading intensity and (**c**) appendicular muscle subgroups. *p*-values refer to the standardized effect sizes. Numbers identify the individual outcomes and are the same in all panels and time points; a number is missing where the outcome was not reported. For Rudroff et al. [63], SDs at 0% TTF were taken from the text (pooled across target forces) because the error bars in the figure could not be assigned unambiguously.

Exploratory meta-regression (bubble plots) indicated that the lower RPE during HIMA at 50% TTF was more pronounced at lower loading intensity (slope +0.018 per % MVIC, *p* = 0.016; **Fig. 5b**), whereas intensity was not associated with the effect at initiation (*p* = 0.167) or at task failure (*p* = 0.602). Within the two studies testing several loads [5, 63], the difference at 50% TTF was likewise smaller at 60% than at 20% MVIC (*g* = −0.36 and −0.11 at 20% vs. +0.23 and +0.35 at 60% MVIC), although not monotonic (45% MVIC: *g* = −0.55) [63]; no consistent pattern was evident at initiation or task failure. Muscle group was not significantly related to the effect at any time point (test of between-subgroup differences: *p* = 0.186, 0.451 and 0.782; **Fig. 5c**), although intensity and muscle group were confounded and outcomes of the same study were treated as independent. Heterogeneity was low at initiation and task failure (I^2^ = 22% and 0%) and moderate at 50% TTF (I^2^ = 60%, Q-test *p* = 0.001).

Considering the significantly shorter TTF for HIMA, the RPE increase rate was higher for HIMA in 14 of 16 outcomes (mean relative difference +51%; **Fig. 6**), nine of which were reported as significant in the primary studies (**Table 1**). This pattern was reinforced by two further studies reporting faster RPE increases for HIMA over a set time [76, 87]. The two outcomes favoring PIMA were non-significant in the primary studies (**Fig. 6, Table 1**).

**Fig. 6.**
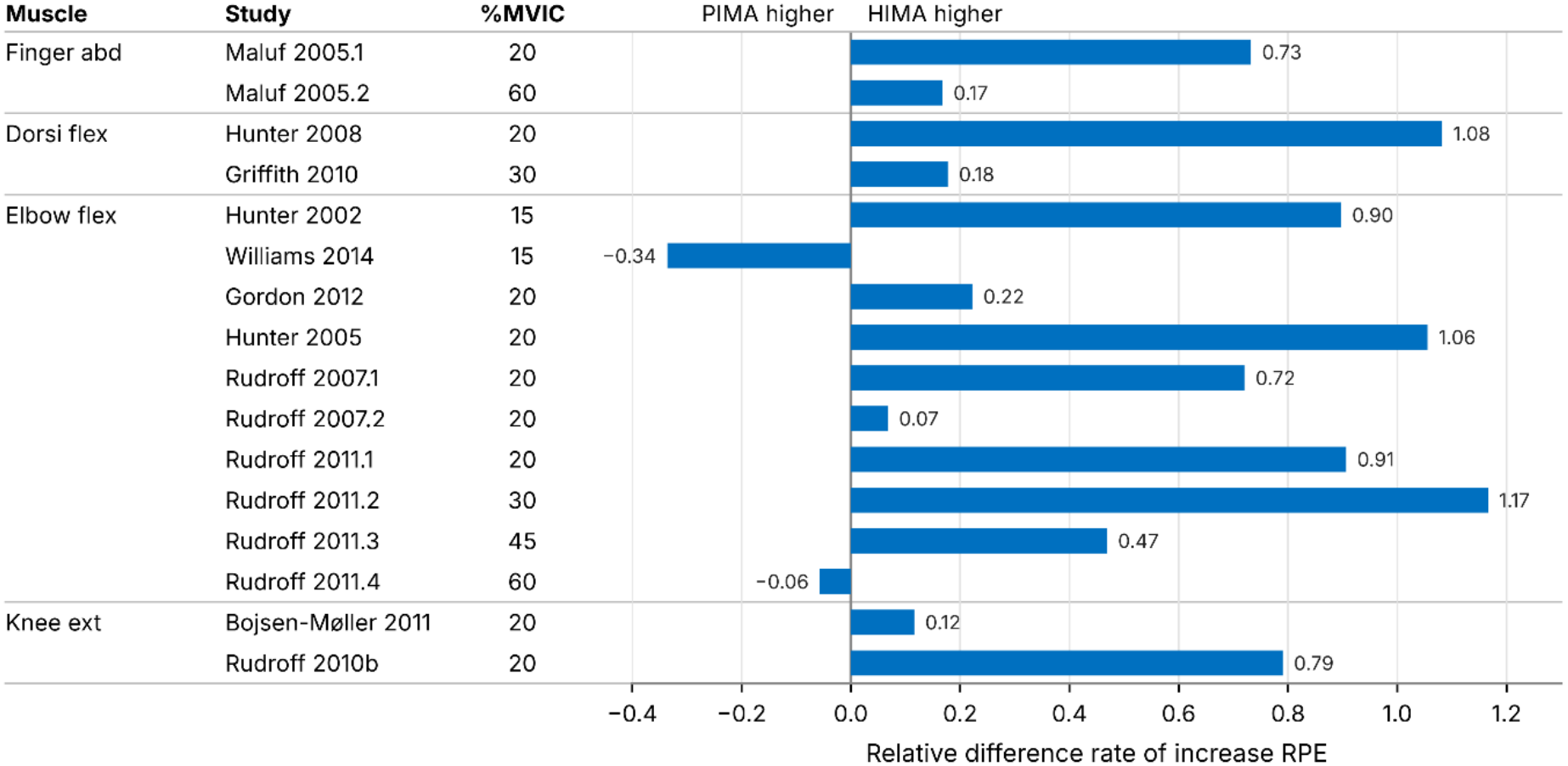
Relative difference in the rate of increase in ratings of perceived exertion (RPE) between HIMA and PIMA for each outcome. Positive values indicate a faster increase during HIMA, negative values during PIMA. *Note:* For each outcome and each task, the rate of increase was calculated as (value at task failure − value at the start of the task) / time to task failure (TTF). The relative difference was calculated as (rate HIMA − rate PIMA) / rate PIMA.

### Neural and neuromuscular parameters

The 45 studies examining neural or neuromuscular differences between PIMA and HIMA are detailed in **Online Resource 4** [3-11, 26, 31, 43-46, 50-52, 54, 63, 64, 69-72, 74-93]. Unlike the mechanical (above) and metabolic (below) variables, more than minimal pooling of findings via meta-analyses or otherwise was impossible due to the exceptional variety of collection methods and data processing. Nearly all studies (n = 41) employed surface electromyography (sEMG) [3-5, 7, 8, 11, 31, 43-46, 50-52, 54, 63, 64, 69-72, 74-93], while five included needle EMG [6, 31, 52, 87, 90], 12 employed intramuscular EMG [4, 5, 44, 50, 64, 70, 75, 76, 78, 85, 87, 92], and three used MMG and mechanotendography (MTG), which mirror oscillatory central activity [9, 10, 26]. Three studies assessed neural function via electroencephalography (EEG) [26, 45, 88]. Twelve studies included peripheral nerve stimulation (electrical, mechanical or magnetic) [3, 11, 43, 46, 51, 54, 69-71, 82, 83, 85], while four studies employed transcranial magnetic stimulation (TMS) [3, 54, 82, 83]. The diverse data processing methods and outcomes included EMG or MMG amplitudes [4, 7-10, 43, 45, 50, 63, 64, 69-72, 74-76, 78, 83, 87, 90-92], root-mean-square (RMS) [5, 44, 46, 54, 77, 79-81, 88, 89, 93], frequency/power [6, 9, 10, 26, 46, 63, 81, 89, 91, 93], burst rates [4, 77-79], burst durations [4, 78], discharge rates [6, 31, 44, 52, 72, 76, 87, 90], ISI CV [31, 72, 76, 90], recruitment rates and thresholds [31, 72, 80, 87], H-reflex [83], heteronymous facilitation [43, 51, 69-71, 82, 85], D1-inhibition [43, 51, 69, 71], D2-inhibition [51], T-reflex amplitude [70], silent period [3, 54, 82, 83], and coactivation ratios [7, 8, 43, 63, 71, 75, 77, 79, 86, 90, 91]. Three studies investigated the coherence of muscle and brain activity [26, 45, 88]. Thirty-one studies examined antagonist muscle groups [3, 5, 7, 8, 31, 43-45, 50, 51, 63, 64, 70-72, 75-81, 83, 85-92], while 34 assessed synergists [3-5, 7-10, 26, 43-45, 50, 51, 54, 63, 64, 69-72, 75-79, 82, 85, 87-93]. Four studies had specific focuses, including injection-induced experimental joint pain [44, 45], residual force enhancement [86], and skin and core temperature manipulation [46]. Given this heterogeneity, and for several parameters, the small number of studies, the following comparisons are descriptive and hypothesis-generating rather than robust effects.

### Neuromuscular activity

EMG amplitude differences (aEMG or RMS) for agonist muscles were significant in only 13 of 48 documented outcomes [4, 5, 7, 8, 43, 50, 54, 63, 64, 69-72, 74-81, 83, 88-93]; five with higher agonist activation during PIMA [4, 63, 64, 78, 83], and eight being higher for HIMA [50, 72, 77, 92]. Where standardized effect sizes were available, they were distributed in both directions: four favoring PIMA (*d* = −6.79 to −0.66, −38.5 to −25.5%) [4, 64, 78, 83] and three favoring HIMA (*d* = 0.22–0.58, 11.8–25%) [77, 92], indicating no consistent direction for steady-state activation. The rate of EMG increase, however, was more consistent: eight of 17 outcomes [4, 5, 54, 63, 72, 77-79, 81, 83, 90, 91] showed a significantly greater increase of agonist EMG amplitude during HIMA [5, 63, 72, 77, 79, 91] and none during PIMA; all six outcomes for which an effect size could be derived favored HIMA (*d* = 0.46– 0.99, 0.1–3566%) [63, 72, 77, 79, 91]. A similar picture emerged for the EMG amplitude of synergist muscles: only four of 15 outcomes [4, 8, 50, 63, 70, 72, 75, 79, 93] were significant— two greater during HIMA (*d* = 0.42–0.72, 22.8–41.4%; reported in the same study) [63] and two greater during PIMA [50, 79], of which one effect size was available (*d* = −0.46, −21.6%) [79]. The increase in synergist EMG [4, 43, 63, 72, 77, 79], however, was greater during HIMA in five of 10 outcomes [4, 63, 77, 79], the remaining five being non-significant; both outcomes with a computable effect size favored HIMA (*d* = 0.43–3.16, 0.5–81%) [63, 79]. For antagonist muscles, only one of 17 relevant outcomes [7, 8, 43, 69-72, 75, 76, 80, 81, 83, 88-92] was significant, with greater EMG amplitude for PIMA [89] (no effect sizes available). The rate of antagonist EMG increase, however, was greater during HIMA in four of nine outcomes [5, 72, 77-79, 81, 83], with all three computable effect sizes favoring HIMA (*d* = 0.40–0.82, 63– 600%) [77, 79, 81].

For agonist EMG burst rate seven outcomes were reported [4, 77-79]. The overall burst rate was higher during HIMA in two of four outcomes (*d* = 0.20–0.69, 44.2–210.5%) [78, 79]; its increase was significantly greater during HIMA in all three reported outcomes (two effect sizes available: *d* = 0.55–0.61, 35.7–40.7%) [77-79]. One study considered the antagonist burst rate, without significant between-task differences [78]. Discharge rates were lower for HIMA in eight of 12 outcomes [31, 52, 72, 76, 87, 90] with seven effect sizes available (*d* = −0.37 to −4.56, −28 to −9.7%) [31, 72, 76, 87, 90]; the remaining four were non-significant (e.g., in older participants or for neutral forearm position) [31, 52, 87, 90]. Discharge rates declined significantly more during HIMA in all three reported outcomes (one effect size: *d* = 0.55, 60%) [76, 87]. The absolute ISI CV showed no consistent pattern across four outcomes—greater during HIMA in two (*d* = 0.57 and 0.80, 23.9% and 46%) [31, 76], lower in one (young participants at large target-force differences, *d* = −0.64, −20.3%) and non-significant in older participants [31]. In contrast, the increase in ISI CV was greater in all three reported outcomes (two effect sizes: *d* = 0.65–2.31, 210–625%) [31, 72, 90].

The H-reflex was examined as a between-task outcome in only one study, where a faster and stronger decline during HIMA was reported (*d* = 1.35, 72%) [83]. Heteronymous facilitation was greater for HIMA in eight of 12 outcomes (seven effect sizes available: *d* = 0.21–1.32, 5.2–34.6%) [51, 69-71, 82, 85], and similar across tasks for older adults (*d* = 0.05) [71] and for mechanical stimulation (no effect size) [85]. One study investigated the heteronymous facilitation over time until failure and found no difference at 2 min (*p* > 0.05), but a progressively decreasing facilitation for HIMA toward task failure (*p* = 0.049; no effect sizes available)—reflecting a more pronounced time-dependent modulation of the conditioned H reflex during HIMA (*p* = 0.02) [43]. D1 inhibition was lower for HIMA in five of seven outcomes (*d* = −0.71 to −3.6, −14.3 to −34.4%) [43, 51, 69, 71]. This pattern held across the relatively short contraction duration (<60 s) of the studies and, in the one study that tracked a fatiguing contraction, was still present at 2 min (*d* = −1.59, −33%); only at task failure did it reverse to greater D1 inhibition during HIMA (*d* = 1.57) [43]—a phase the other studies did not examine. The remaining outcome without a difference across tasks was reported for old participants (*p* = 0.63) [71].

For the coactivation ratio, an effect size was available for only one of 12 outcomes [7, 8, 43, 63, 71, 75, 79, 86, 90, 91]; this outcome (wrist extensors of older adults) showed a trivial effect (*d* = 0.15, *p* = 0.02) [71]. The remaining 11 outcomes did not differ significantly between muscle actions. The increase of coactivation ratio was significantly greater during HIMA in two outcomes of one study (*p* = 0.01; no effect size) [77].

MMG and MTG—as mechanical counterpart to EMG—were investigated in two studies comparing HIMA and PIMA at high intensities (80–90% MVIC) for elbow extensors, in a single-person and a coupled (interpersonal) setting. MMG/MTG amplitude and frequency over the whole duration were statistically similar between tasks in either study [9],[10]. In the single-person setting, MMG amplitude at the end of the failure task was lower during HIMA (*d* = −0.39, *p* = 0.012), whereas the overall amplitude variation of triceps MMG/MTG (*d* = 0.32–0.54, n.s.) and the power of triceps MTG (*d* = 0.42, *p* = 0.037) were higher for HIMA [9]. In the interpersonal setting—which additionally requires partners to continuously attune to one another—the pattern shifted to the abdominal oblique muscle (stabilizer): higher amplitude variation (15 s task: *d* = 0.71, *p* = 0.013; fatiguing task: *d* = 0.58, *p* = 0.007) and lower power (15 s task: *d* = −0.39, *p* = 0.001; fatiguing task: *d* = −0.28, *p* = 0.011) during HIMA; similarly, the triceps tendon (MTG) showed a non-trivial tendency towards lower power during HIMA (*d* = −0.27 to −0.31, n.s.) [10].

### Brain activity

Three studies included measures of brain activity via EEG [26, 45, 88]. Poortvliet et al. [45, 88] investigated the knee extensors (10% MVIC, 30 s) and reported non-significant differences between tasks for corticomuscular coherence (central areas of EEG were used that approximated the motor cortex of the left hemisphere), but significantly lower cortico-cortical coherence for HIMA (four of five outcomes; *p* < 0.001) though with trivial-to-small effect sizes (*d* = −0.09 to −0.27, −5 to −14.5%), and significantly but negligibly higher left-hemisphere beta-band EEG power for HIMA (*p* < 0.05, *d* = 0.11, 0.68%) [88]. Schaefer and Bittmann [26] examined the coherence between EEGs (central, left and right areas), MMGs (triceps brachii and ipsilateral abdominal external oblique muscles; statistics did not consider muscles separately) and force-signal during interpersonal actions of elbow extensors (70% MVIC of the weaker partner, fatiguing, n = 2). Significantly higher values were revealed for HIMA regarding EEG coherence (central and left areas) to the partner’s MMG (inter-brain-muscle coherence), force-EEG-coherence (central and right area) and the frequency of inter-brain-synchrony (central vs. left and central vs. right) and inter-brain-muscle-coherence (EEG right vs. MMG), respectively (*p* ≤ 0.047, *d* = 0.36–2.67, 5.1–67.6%) [26].

### Special factors

Because of their distinct experimental paradigms, the following studies are described narratively; where a paired between-task comparison was precluded, the corresponding data were not included in the pooled outcome counts (Table 1). Coletta et al. [46] reported greater RMS of the EMG signal for HIMA at modest heating and cooling conditions (±0.5 °C) during 3 s muscle actions. However, non-significant muscle action differences were found during 60 s muscle actions at the higher heating (1 °C) condition [46]. It must be noted that the RMS was significantly higher for HIMA at baseline. Marion and Power [86] compared PIMA and HIMA following active lengthening of the dorsi-flexors to examine residual force enhancement, with no significant differences between isometric tasks for any outcome (activation reduction, neuromuscular economy, coactivation).

Experiments led by Poortvliet investigated the effects of hypertonic saline injections into the infrapatellar fat pad to temporarily generate knee pain during low-intensity (∼10%) knee extensor muscle actions [44, 45]. The first study considered 35 single-motor-units (n = 5), which were assessed across all conditions (no pain, pain, HIMA, PIMA) in a single session [44]. Additionally, 189 single-motor-units (n = 11) were assessed over two sessions. For the 35 single-motor-units, the no-pain state showed similar RMS and discharge rates between HIMA and PIMA, where the discharge rate variability tended to be higher for PIMA (*p* = 0.05, *d* = −0.2). Under pain, the decline in discharge rate was greater for PIMA. Furthermore, discharge rate variability between no-pain and pain states only changed significantly for PIMA. Considering all 189 single-motor-units, discharge rate was significantly higher for HIMA across pain states (main effect). However, discharge rate decreased under pain for both tasks, with a trend towards lower discharge rates for PIMA. Moreover, the number of single-motor-unit discharge rates that changed (>10% increase or decrease) from the no-pain to the pain state was twice as high in PIMA (44.9%) as in HIMA (22.2%) [44]. The second study by Poortvliet et al. examined the effect of pain on corticomuscular coherence: only PIMA showed significant changes between no-pain and pain states (lower coherence under pain), together with decreased beta- and increased gamma-band power; the authors suggested that brain activity is less affected by pain during HIMA [45].

### Cardiovascular and metabolic parameters

Fourteen studies examining cardiovascular or metabolic differences between PIMA and HIMA are detailed in **Online Resource 5** [4, 5, 7, 8, 47, 63, 64, 75, 77-79, 87, 91, 92]. Eleven studies assessed HR and MAP [4, 5, 63, 64, 75, 77-79, 87, 91, 92]: seven utilized the elbow flexors [4, 63, 64, 75, 78, 87, 92], while others employed the dorsi-flexors [77, 79], knee extensors [91], or first dorsal interossei [5]. Two studies compared parameters of muscle oxygenation using near-infrared spectroscopy on biceps and triceps brachii during elbow flexions at 20% or 60% MVIC [8], or an O2C^®^ spectrophotometer on biceps brachii during elbow flexions at 60% MVIC [47], with both studies reporting non-significant between-task differences. Another study used positron emission tomography/computed tomography (PET/CT) with intravenously administered [^18^F]-fluorodeoxyglucose to quantify tracer uptake (standardized uptake values, as an index of glucose uptake) in the agonist (knee extensors), antagonist, and other lower-extremity muscles at 25% MVIC [7]. In older adults, all between-task differences were non-significant and trivial-to-small (*p* > 0.05, *d* = −0.13 to −0.26, −7.7 to −4.3%). In young participants, glucose uptake was greater during HIMA, with non-trivial effects for the agonist (*p* < 0.01, *d* = 0.68, 32.6%) and antagonist (*p* < 0.05, *d* = 0.47, 15.4%) but only a trivial effect for hip muscles (*p* < 0.01, *d* = 0.19, 9.7%) [7].

Nine studies allowed meta-analysis for HR and MAP [4, 5, 63, 64, 75, 77-79, 91], whereas two studies terminated both tasks after a predefined duration rather than at task failure and were therefore not included [6, 92]. Meta-analyses of 10–14 outcomes (6–9 studies; 28.6 ± 15.2% MVIC, range: 15–60%) found trivial differences in HR and MAP at initiation, 50% TTF and at task failure (all |*g*| < 0.20; **Fig. 7a, 8a**) [4, 5, 63, 64, 75, 77-79, 91]. For HR (n = 116–172), values were slightly higher during HIMA at initiation (g = 0.15 [0.01, 0.29], *p* = 0.030), with no significant differences at 50% TTF (*g* = −0.14 [−0.34, +0.05], *p* = 0.154) or at task failure (*g* = 0.00 [−0.14, +0.15], *p* = 0.984). For MAP (n = 132–172), no significant differences were found at initiation (*g* = 0.11 [−0.10, +0.32], *p* = 0.285) or at 50% TTF (*g* = 0.05 [−0.14, +0.24], *p* = 0.600), whereas it was slightly higher during HIMA at task failure (*g =* 0.19 [0.05, 0.32], *p* = 0.008). Sensitivity analyses with different assumed within-participant correlations (*r* = 0, 0.3, 0.5, 0.7) showed that the HR difference at initiation was only significant for *r* = 0.5 and 0.7, but not for *r* ≤ 0.3, whereas the MAP difference at task failure was significant for *r* ≥ 0.3 (**Online Resource 6**).

**Fig. 7.**
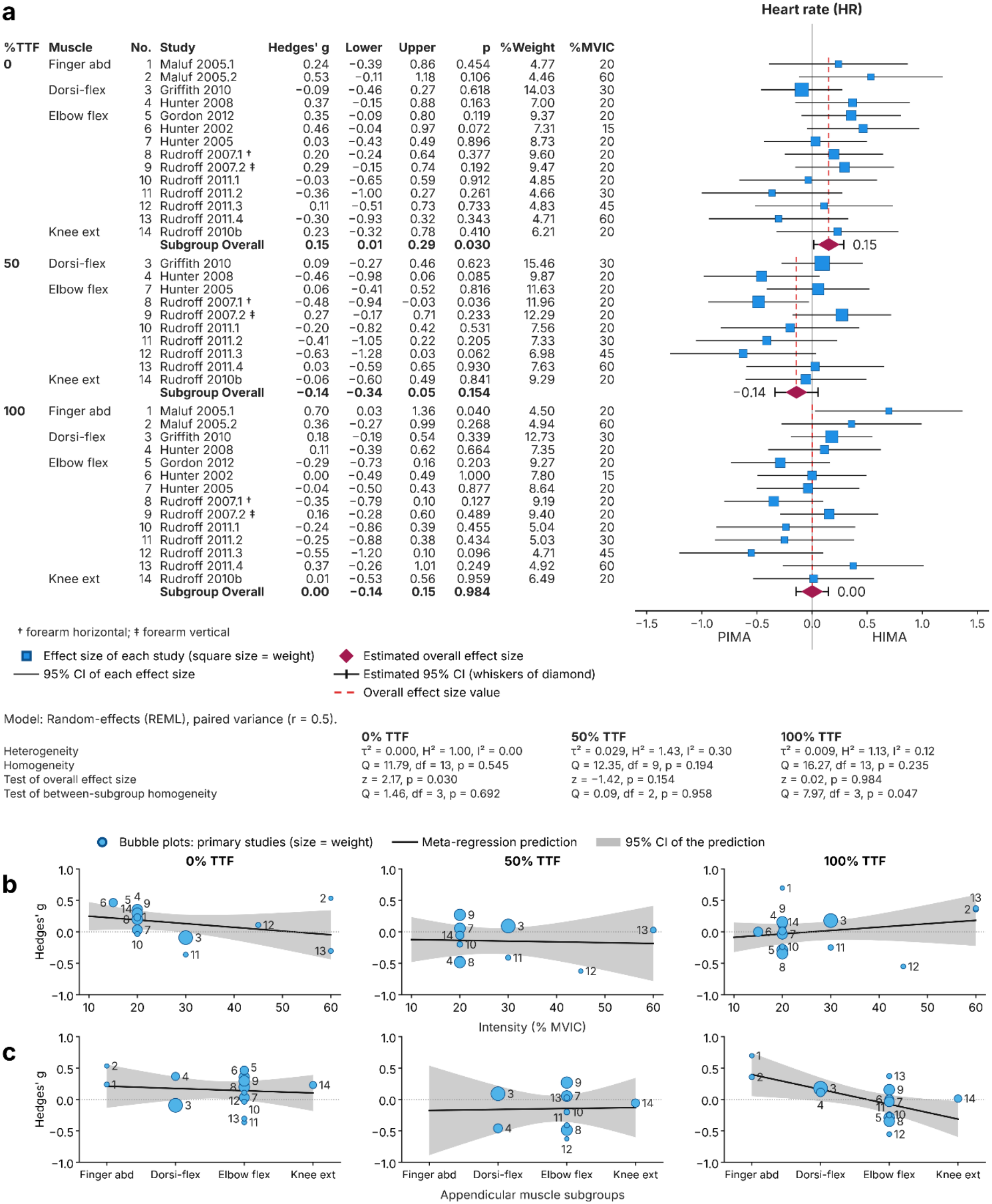
Meta-analysis of heart rate (HR) in PIMA vs. HIMA at 0%, 50% and 100% of time to task failure (TTF). (**a**) Forest plot of Hedges’ g (negative values: lower HR during HIMA); random-effects model with paired variance. Bubble chart regressions for (**b**) loading intensity and (**c**) appendicular muscle subgroups. *p*-values refer to the standardized effect sizes. Numbers identify the individual outcomes and are the same in all panels and time points; a number is missing where the outcome was not reported.

Examining bubble plots is complicated by relatively few studies. Exploratory meta-regression revealed no association with loading intensity (all *p* ≥ 0.288), although intensity and muscle group were confounded and outcomes from the same study were treated as independent. HR at task failure differed between muscle groups (*p* = 0.047): it tended to be higher for PIMA for larger muscles, with the reverse for smaller muscle groups (slope −0.24 per muscle-group rank, *p* = 0.014; **Fig. 7c**). The opposite appeared true for MAP during the initial phases (0% and 50% of TTF): the larger the muscle group, the more HIMA seemed to elevate MAP compared to PIMA, although not significantly (*p* = 0.119–0.146; **Fig. 8c**). Heterogeneity was low for HR (I^2^ = 0–30%) and low to moderate for MAP (I^2^ = 0–55%, highest at initiation, I^2^ = 55%, Q-test *p* = 0.005).

**Fig. 8.**
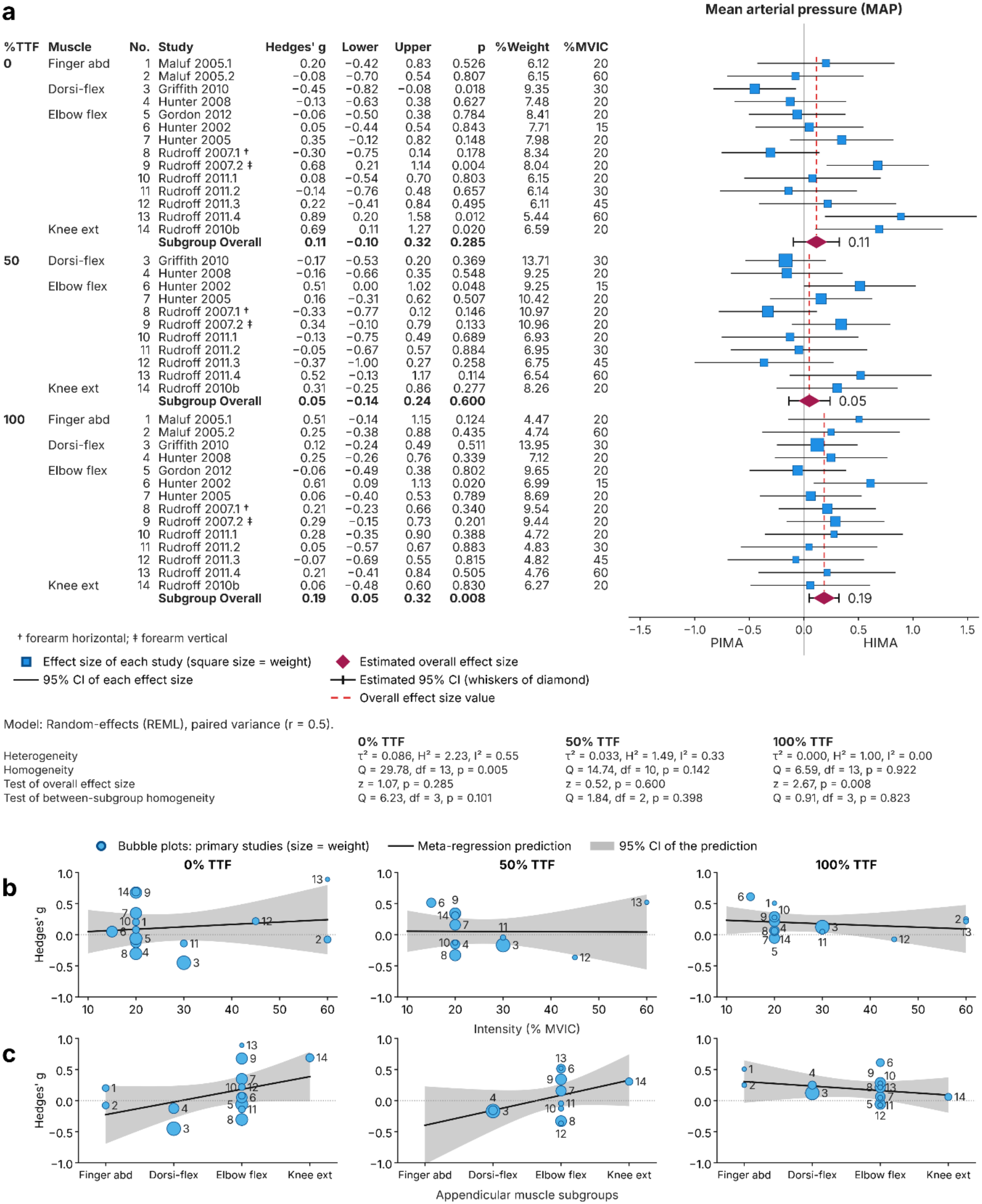
Meta-analysis of mean arterial pressure (MAP) in PIMA vs. HIMA at 0%, 50% and 100% of time to task failure (TTF). (**a**) Forest plot of Hedges’ g (negative values: lower MAP during HIMA); random-effects model with paired variance. Bubble chart regressions for (**b**) loading intensity and (**c**) appendicular muscle subgroups. *p*-values refer to the standardized effect sizes. Numbers identify the individual outcomes and are the same in all panels and time points; a number is missing where the outcome was not reported.

The rate of HR and MAP increase was higher during HIMA in most outcomes (HR: 10 of 14, mean relative difference +52%; MAP: 12 of 14, +77%; **Fig. 9**) [4, 5, 63, 64, 75, 77-79, 91]. Because TTF was mostly shorter during HIMA, this indicates a faster cardiovascular response relative to time rather than a greater absolute response.

**Fig. 9.**
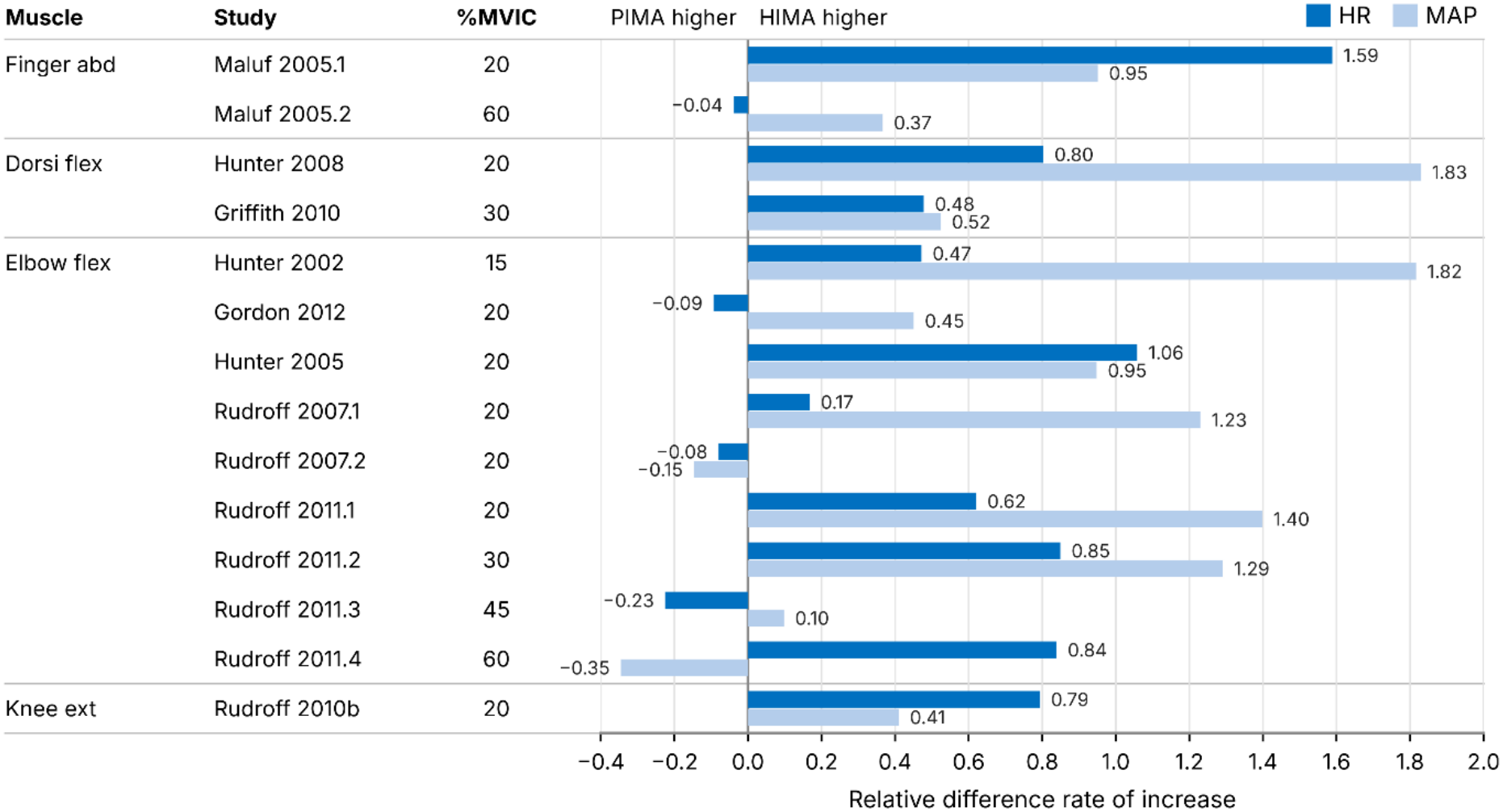
Relative difference in the rate of increase in heart rate (HR; dark blue) and mean arterial pressure (MAP; light blue) between HIMA and PIMA for each outcome. Positive values indicate a faster increase during HIMA; negative values indicate a faster increase during PIMA. *Note:* For each outcome and each task, the rate of increase of HR or MAP, respectively, was calculated as (value at task failure − value at the start of the task) / time to task failure (TTF). The relative difference was calculated as (rate HIMA − rate PIMA) / rate PIMA.

## Discussion

In sports science and medicine, it is uncommon to differentiate the two isometric muscle action types. PIMA (the most common isometric muscle action) is usually assessed to quantify muscle strength. However, focus is increasingly shifting towards HIMA, which is being utilized more frequently by sports medicine and performance practitioners aiming to improve musculotendinous morphology and neuromuscular function [38-42]. Despite a wealth of studies comparing PIMA and HIMA, it is difficult for researchers and practitioners to understand their potential applications due to the wide range of methods and data reporting. Therefore, we identified and presented all studies directly comparing the isometric muscle action types. The first question was whether holding and pushing isometric muscle actions can be distinguished experimentally. Based on the available evidence, the answer appears to be *yes*, with HIMA and PIMA differing across several objective measures. When examining the 54 studies comparing both isometric types, our findings can be separated into those supported by meta-analysis and those derived from structured narrative synthesis. First, meta-analytic evidence demonstrated that (1) PIMA leads to longer TTF at the same absolute intensity (except for postural axial muscles), although this difference may narrow at higher intensities; (2) differences in HR and MAP at matched relative time points were trivial (|*g*| < 0.20), including two significant differences (slightly higher HR at task initiation and MAP at task failure during HIMA); and (3) RPE did not differ at task initiation and failure—with the exception of a higher RPE during PIMA at 50% of TTF (*g* = −0.31). Second, structured narrative synthesis suggested several recurring patterns. PIMA was generally associated with larger force fluctuations, higher discharge rates, greater D1-inhibition, and higher peak torques. HIMA tended to show higher heteronymous facilitation, greater EMG burst rates, greater glucose uptake in young participants, and faster increases in force or position fluctuations, EMG amplitude, ISI CV, RPE, HR, and MAP. In contrast, no consistent direction emerged for the absolute ISI CV. For MVIC decline, effect sizes were evenly distributed between directions, although the few significant outcomes consistently indicated a greater decline for HIMA. No consistent differences (largely non-significant) emerged for absolute EMG amplitude and frequency, coactivation, motor evoked potentials (MEP), and oxygenation. Finally, several individual studies reported differences between HIMA and PIMA in cortico-cortical and corticomuscular coherence, as well as differential responses to experimentally induced joint pain. These narrative findings should be interpreted cautiously, as most variables beyond TTF, RPE, HR, and MAP were not amenable to meta-analysis and often relied on single-study comparisons or heterogeneous methodologies.

### Distinguishing holding and pushing isometric muscle actions

#### Muscle fatigability

TTF at the same intensity was the most frequently investigated parameter, resulting in the clearest findings. TTF was significantly longer in appendicular muscles for PIMA, with a large meta-analytic effect (*g* = −0.89). These findings suggest that one or more factors must contribute to reduced endurance with HIMA. The effect was larger at low intensities (≤30% MVIC) than at high intensities (≥50% MVIC; *g* = −1.02 vs. −0.45), suggesting that the difference may be attenuated with increasing intensity [5, 8, 63], although meta-regression showed no linear association and intensity was confounded with laboratory. Only six studies investigated TTF with intensities ≥50% MVIC [5, 8, 9, 47, 63, 81]; four outcomes showed small to large effects for shorter TTF during HIMA (*g* = −1.16 to −0.35; three significant in meta-analysis, **Fig. 4a**) [8, 9, 25, 81] and two were trivial [5, 63]. The largest effect at high intensity (*g* = −1.16, 80% of MVIC) was found by Schaefer et al. [9], who used a pneumatic device to exert an external force during the holding task against the participant without permitted position corrections. In contrast, other studies employed inertial loads, with four explicitly allowing position correction [5, 8, 63, 81]; in these, effects were trivial to moderate (*g* = −0.54 to 0.07; three non-significant in meta-analysis). It is suggested that minor muscle shortenings during these position corrections might alter neuromuscular control processes in favor of PIMA. Dech et al. [47] support this, as dumbbell-loaded HIMA were examined without and with brief, subtle muscle twitches at 60% MVIC. TTF during HIMA with twitches was larger than without and did not differ from PIMA [47]. However, some studies investigating low intensities also allowed position corrections during HIMA, finding moderate to large differences (all significant in meta-analysis) [5, 8, 43, 63, 72, 75]. Consequently, intensity could play a role in whether minor muscle shortenings can alter neuromuscular control. Differences between the isometric types, even at high intensities, are further supported by the investigation during paired measurements (90% MVIC) [10]. In most cases (85%), the partner performing HIMA failed first. As the settings for HIMA and PIMA were identical (load compliance, i.e., position stability and correction possibilities), the differences must have resulted from the assigned tasks.

Axial muscle TTF was significantly longer during HIMA [53, 93]—the only subgroup with an effect in the opposite direction, whereas differences among the appendicular subgroups were minimal. This likely reflects different habitual neuromuscular functions compared with appendicular muscle groups. Axial muscles are routinely engaged in stabilizing and “holding” the torso during standing, walking, and upright sitting, and therefore habitually perform tasks analogous to HIMA. In contrast, appendicular muscles more commonly execute voluntary movements resembling PIMA. In addition, during axial muscle testing, participants must support not only the external load, but also a substantial portion of their body mass, which may have caused the actual intensity experienced to exceed the nominal target. Although the available results are limited to two studies, these patterns suggest that axial and appendicular muscles may need to be considered separately in both assessment and treatment contexts.

The significant TTF differences between HIMA and PIMA suggest that, at least for appendicular muscles, HIMA is more challenging to maintain than PIMA. One factor might be the relative proportion of peak force achieved via each muscle action. The intensity in nearly all studies was based on PIMA_max_ (MVIC). HIMA_max_ has been estimated to be ∼77% of PIMA_max_ [21, 22, 25]. Thus, the absolute loads were identical, but the relative loading presumably was not. Fluctuations in force and/or position tended to increase more rapidly during HIMA compared to PIMA, whereas absolute fluctuations were generally greater during PIMA. Similar findings for TTF and force decrements revealed no substantial intermuscular differences, suggesting minimal influence of total muscle mass or limb region. However, the relatively small number of studies and outcome measures limits the ability to draw definitive conclusions about between-muscle differences. Overall, the question remains about why lower peak forces and shorter TTF occur during HIMA.

#### Neurophysiological, metabolic, and cardiovascular considerations

In line with shorter TTF, several parameters increased faster during HIMA (e.g., fluctuations, RPE, MAP, HR, EMG amplitude, burst rate), suggesting that HIMA is more strenuous than PIMA. Since perceived effort and MAP are influenced by central processes [77, 94], these findings are consistent with the hypothesis that HIMA requires greater central processing. The two studies comparing muscle oxygenation found no statistically significant task differences (where available, trivial to small effects: *d* = −0.08 and −0.24) [8, 47], which argues against a predominantly metabolic explanation. Studies suggest that HIMA involves greater supraspinal and spinal activation compared to PIMA, as indicated by, for example, spinal processing reflected in earlier and greater changes in motor-unit recruitment and rate coding during HIMA [4, 5, 63, 72, 79, 80, 87, 91]. Moreover, the lower D1-inhibition and higher heteronymous facilitation during HIMA indicate reduced presynaptic inhibition and enhanced reliance on proprioceptive input [43, 51, 69-71, 82, 85]. Notably, this pattern was not static. In the only study tracking a fatiguing contraction to task failure, presynaptic inhibition of Ia afferents was reduced early during HIMA but not during PIMA, whereas at task failure the tasks diverged in the opposite direction, with greater inhibition during HIMA (task × time, *p* < 0.001) [43]. Functionally, an early reduction increases the spindle feedback reaching the motor neuron pool—plausibly relevant when joint position must be monitored and corrected continuously, as during HIMA. The late increase would have the opposite effect: with stronger presynaptic inhibition, less spindle input reaches the motor neurons, reducing their excitatory drive and slowing their discharge. This aligns with the present synthesis—discharge rates were lower during HIMA than PIMA in eight of 12 outcomes (seven with non-trivial effect size) [31, 72, 76, 87, 90]—suggesting a coherent chain from afferent gating to motor output. A progressive withdrawal of afferent support towards task failure could thus contribute to the shorter TTF during HIMA. As only one study has tracked Ia gating across a fatiguing contraction to task failure, this should be regarded as a hypothesis for targeted testing rather than an established mechanism. Presynaptic inhibition is itself primarily controlled by supraspinal centers and sensory feedback [95], reinforcing the relevance of supraspinal pathways when distinguishing between isometric tasks. Accordingly, a strategy for HIMA was suggested “in which supraspinal centers choose to enhance the contribution of muscle afferents to the synaptic input that converges onto spinal motor neurons” [71, 96], aligning with findings that HIMA requires greater proprioceptive information [11, 82, 87]. Taken together, these findings support the hypothesis, proposed by several researchers, that HIMA involves different, potentially more complex central modulation and sensorimotor integration than PIMA [3, 5, 11, 12, 26, 31, 63, 64, 69, 75, 78, 80, 87, 92].

The greater complexity is often attributed to the higher instability of the HIMA setting, which requires greater reflex responsiveness [69]. Indeed, 46 of 54 studies used inertial loading, or similar methods, to realize HIMA, allowing movement across muscle shortening and lengthening. In contrast, four studies employed a pneumatic device that impedes muscle shortening [9, 21, 22, 25], while another four utilized personal interaction [10, 23, 24, 26]. In those studies, the ‘stability’ or ‘freedom-to-move’ was similar between tasks, yet differences between HIMA and PIMA were still observed. Thus, movement freedom alone cannot explain all task differences.

While load compliance (i.e., force vs. position control) plays a major role in shaping neuromuscular demands, it is important to clarify that HIMA and PIMA are not synonymous with closed- and open-chain kinetic exercises. Both HIMA and PIMA can be performed under either chain condition, depending on joint fixation, limb support, or load configuration. The key distinction lies in whether the individual is actively generating force against an immovable object (PIMA) or reactively stabilizing against an imposed force (HIMA), regardless of limb constraint. Equating these with closed- or open-chain exercises risks oversimplifying their neural and sensorimotor distinctions, which are central to this review. Overall, the available evidence suggests that differences in neural control contribute to the distinct responses observed between PIMA and HIMA, although their contribution relative to task mechanics and experimental configuration remains to be determined.

#### Neural mechanisms during HIMA and PIMA

The meta-analytic finding of shorter TTF during HIMA raises the question of underlying neural mechanisms. Available evidence suggests differences at both supraspinal and spinal levels. At the supraspinal level, Poortvliet et al. [88] reported significantly lower cortico-cortical coherence (*d* = −0.09 to −0.27) but higher beta band power (*d* = 0.11) during HIMA, which they interpreted as reflecting greater subcortical involvement; however, these effects were only trivial to small. In a case study of two coupled participants, higher coherence between the partners’ cortical and muscle activity was found for HIMA, with small to large effects (*d* = 0.36–2.67) [26]. Despite considerable methodological differences between studies, both point towards differences in cortical processing between the isometric types, though this evidence remains preliminary.

At the spinal level, the task- and time-dependent modulation of presynaptic inhibition described above provides a direct interface between the two levels: presynaptic inhibition is set by descending pathways yet acts on afferent transmission to the motor neuron pool, thereby determining how much proprioceptive information is available for control. These differences may therefore be linked to distinct roles of afferent input. A potential paradox arises from Poortvliet et al.’s [44, 45] finding that experimental pain produced greater motor modifications during PIMA than HIMA—seemingly inconsistent with HIMA involving heightened sensory processing [4, 5, 44, 64, 79, 89]. Yunoki et al. [11] proposed that sensory input unrelated to movement is suppressed during PIMA (especially cutaneous information), while HIMA maintains greater openness to proprioceptive feedback. Under this framework, HIMA would be selectively more responsive to proprioceptive input than PIMA. In support of this interpretation, sensorimotor modulation was more pronounced during HIMA [6, 82, 87], and interspike interval variability increased more rapidly [31, 72, 90], indicating enhanced feedback-dependent regulation.

Collectively, these findings suggest that HIMA relies on continuous proprioceptive updating integrated across supraspinal and spinal levels, consistent with models of optimal feedback control [96, 97]. The persistence of differences in task failure even in paired settings with identical mechanical conditions and high target force [10] reinforces that these differences are not solely attributable to the mechanical configuration or task artifacts; rather they may reflect genuine differences in control strategy. If confirmed by future studies, these findings suggest that distinguishing between HIMA and PIMA may be relevant when classifying isometric tasks and designing diagnostic or training protocols. In particular, HIMA may serve as a more sensitive probe of sensorimotor integration capacity. Confirmation through further research with standardized protocols is needed.

#### HIMA and PIMA as ‘stopped’ or ‘restricted’ lengthening or shortening actions

Classically, isometric muscle action is considered a single category, defined by constant muscle length. However, our findings demonstrate that HIMA and PIMA differ in several parameters, challenging the notion of uniform isometric muscle action. Moreover, even during isometric tasks, fascicles shorten slightly while tendons elongate [98, 99], and mechanical oscillations reflect that muscles continuously alternate between shortening and lengthening [9, 10, 100], which are similar to those during concentric and eccentric actions [100]. These observations have led to the proposal of the ‘stopped-action hypothesis’, inspired by Garner and colleagues [74] and proposed by members of the present authorship team [9, 10].

During PIMA, the participant pushes against immovable resistance—a concentric action that is mechanically prevented from producing movement. During HIMA, the participant resists an external load that would cause lengthening if force were insufficient— also known as eccentric quasi-isometric muscle action [15-19]—resembling a restricted eccentric action. This framework has explanatory value because lengthening muscle actions are known to require more complex neural control strategies than shortening ones [101]. If HIMA indeed resembles a restricted eccentric action, this could account for the proposed differences in motor control strategies between HIMA and PIMA [9-12, 22-24, 26-29, 64]. However, direct comparison between isometric and anisometric actions is complicated by the fact that the latter involves macro changes in muscle length and joint movement.

Despite this limitation, some similarities exist between the isometric types and their suggested anisometric counterparts. Discharge rates are systematically lower during lengthening than shortening actions [101]; similarly, discharge rates were lower during HIMA than PIMA in eight of 12 outcomes [31, 72, 76, 87, 90]. The faster RPE increase during HIMA [4, 5, 63, 64, 73, 75-79, 87, 91] parallels the force overestimation reported for eccentric vs. concentric actions [94]. Modulations of neural activation during lengthening actions are assumed to involve supraspinal and spinal mechanisms, predominantly mediated by spinal pre- and postsynaptic inhibitory mechanisms [101]. Similar mechanisms have been suggested for HIMA [4, 5, 43, 51, 63, 64, 69-72, 76, 79, 80, 82, 83, 85, 87, 91]. Higher central activity (EEG) accompanies reduced EMG activity during lengthening vs. shortening actions [102]; similarly, significantly higher beta band power was found for HIMA [88], though the effect was negligible and data remain limited. Finally, voluntary activation is lower during maximal lengthening than shortening actions, limiting the realization of the muscle’s intrinsic force capacity [101]. A similar mechanism may explain the lower peak torque for HIMA compared to PIMA [21, 22, 25]. H-reflex data are insufficient for comparison, as only one study examined it as a between-task outcome, reporting a faster and stronger reduction during HIMA vs. PIMA [83]; despite this limited evidence, the finding aligns with the typically depressed H-reflex during active and passive lengthening actions [94, 101].

However, some findings are inconsistent with the hypothesis above. MEP and post-TMS silent period are reduced and briefer during lengthening vs. shortening actions [101], but HIMA vs. PIMA results are mixed: MEP differences were non-significant [3, 82, 83], and silent periods were briefer [82], non-significant [54, 83], or longer [3].

In summary, several findings synthesized in the present review are consistent with this hypothesis—including differences in time-to-task failure, motor unit behavior, and selected neurophysiological measures—whereas others remain inconsistent or insufficient. Alternative explanations related to task mechanics, experimental configuration, or differences in relative loading cannot presently be excluded. Further targeted research is required.

#### HIMA with variable external forces – A greater challenge for motor control

Research on Adaptive Force (AF), introduced and to date investigated by members of the present authorship team, provides further observations consistent with the hypothesis of more complex control strategies during HIMA. AF refers to the neuromuscular capacity to adapt to increasing external loads and is assessed via HIMA with a non-stationary load that ramps to and beyond HIMA_max_ [25, 27]. This task requires constant sensorimotor updating [103]. As previously mentioned, adaptive HIMA_max_ (i.e., maximum isometric AF) was significantly lower than PIMA_max_ [21, 22, 25]. Moreover, adaptive HIMA_max_ is reduced significantly in healthy participants by stimuli such as negative imagery or odors [27-29] and by muscle spindle slack induced through specific pre-conditioning [23, 24], while maximal force output remained similar or even increased. In patients with knee osteoarthritis, adaptive HIMA_max_ was also reduced [21]. Whether PIMA_max_ would react similarly has not been directly tested, though findings from Bittmann et al. [24] suggest otherwise. In that study, adaptive HIMA_max_ of the elbow flexors was assessed after two pre-conditioning protocols: (1) muscle action in a lengthened position with passive return to the testing (middle) position (provoking spindle slack), and (2) the same, but followed by a brief voluntary contraction (20% MVIC) in the testing position. Only protocol (1) reduced the adaptive HIMA_max_ significantly (−47%); the brief voluntary contraction in protocol (2) restored full holding capacity. Critically, PIMA inherently begins with voluntary force production, preventing spindle slack from affecting performance. This suggests that HIMA’s reliance on reactive, feedback-dependent control renders it selectively vulnerable to proprioceptive perturbations—a vulnerability likely not shared by PIMA.

These findings suggest that adaptive HIMA is particularly sensitive to afferent input, consistent with the proposed differences in sensorimotor modulation between the two isometric types. This sensitivity may arise from the need to regulate length–tension control while continuously adapting to external loads, which requires the integration of proprioceptive afferents via feedback control mechanisms likely engaging subcortical structures (thalamus, cerebellum, basal ganglia) [104-106]. In PIMA, by contrast, joint position is maintained by the immovable resistance, so that the task itself does not require continuous adaptation of muscle length to the external load, and this integration is presumably less critical. A candidate mechanism is the descending modulation of gamma motor neuron excitability, largely via reticulospinal pathways: gamma drive sets the sensitivity of the muscle spindle itself and thereby the afferent signal available for control [107, 108]. This complements the presynaptic gating discussed above, which determines how much of that signal reaches the motor neuron pool. Both are under descending control, providing two points at which non-motor inputs— nociceptive, autonomic or emotional—could alter the afferent information on which HIMA depends. This fits the spindle-slack findings described above [23, 24], a manipulation acting at the receptor level. Under such perturbations, the neuromuscular system seems to fail to maintain the required position under increasing load, resulting in muscle lengthening.

Consistent with this, adaptive holding capacity was selectively impaired in muscles classified as unstable in semi-professional football players—on average 61% below maximal force output—and discriminated strongly between stability categories (*d* = 6.62), whereas MVIC and the hamstrings-to-quadriceps ratio did not [109]. This indicates that the adaptive holding function can be selectively compromised while maximal force output remains intact— a dissociation that would be invisible to conventional strength testing.

### Potential practical applications

The findings of this review may have implications for injury prevention, diagnostics, rehabilitation and sports performance; however, these are currently based primarily on acute mechanistic evidence and should be considered hypothesis-generating.

#### Injury prevention and diagnostics

Most injuries occur without contact [110] during muscle lengthening, when a muscle must hold or decelerate an external force [111]. Despite this, muscle strength is usually assessed via PIMA, neglecting the adaptive holding component which is closer to the nature of injury mechanisms. One hypothesis arising from the present review is that assessing HIMA— particularly adaptive HIMA—may help identify functional impairments that are not detected by conventional strength testing (see above); this holds potential for diagnostic applications. Experimental studies have demonstrated that adaptive HIMA is adversely affected by proprioceptive disruption, nociceptive input, and negative emotions [21, 23, 24, 27-29, 112]. If comparable reductions occur in real-world settings, they might explain why non-contact injuries can occur suddenly during routine movements under normal load (e.g., running, direction changes, landing) that have been performed thousands of times without incident. Initial evidence is consistent with this hypothesis: in a cross-sectional study of football players (preprint), stability deficits and musculoskeletal complaints were strongly associated, with the ratio of adaptive holding capacity (adaptive HIMA_max_) to maximal force discriminating affected from unaffected players (AUC = 0.93) where maximal strength and the hamstrings- to-quadriceps ratio did not (AUC = 0.41 and 0.43); preliminary follow-up data provide tentative support for a temporal relationship [113]. However, no prospective studies have directly investigated whether reduced holding capacity (muscle instability) increases injury risk or whether preserved holding capacity (muscle stability) is protective. Beyond screening, adaptive HIMA_max_ may also have diagnostic potential: reduced values were found to recover immediately in response to supportive stimuli [112, 114, 115], so its assessment could help identify effective, individualized interventions. Further research is needed to establish clinical utility and to clarify how different stimuli affect HIMA vs. PIMA.

The evidence for isometric exercise in injury prevention is limited. Two low-quality studies suggest reduced injury rates with added isometric training [116, 117], but the type of isometric exercise was not specified. Complex exercises involving reactive stabilization, such as the Nordic hamstring exercise, may be more relevant. Two reviews found significant reductions in hamstring injuries [118, 119], though another reported inconclusive evidence of a protective effect [120]. In a review, Ullman et al. [121] claimed that the positive effects of isometric exercise on injury prevention are assumed because of post-activation potentiation. However, potential distinctions between HIMA and PIMA were not considered.

Collectively, the current evidence supports the generation of two testable hypotheses: (1) adaptive HIMA uncovers functional impairments undetected by conventional strength testing and (2) such impairments are relevant for injury risk. Reductions in adaptive HIMA likely reflect interfering inputs (e.g., nociception, emotional stress) rather than insufficient muscle strength, as they occur immediately after such stimuli, while maximal force output remains similar or increases [23, 24, 27-29]; conventional strength training may therefore not resolve such deficits, which may instead require addressing the interfering inputs themselves. These hypotheses require empirical validation.

#### Rehabilitation

Clinical interest in isometric exercise for conditions like tendinopathy has increased [122], although existing intervention studies generally do not distinguish between PIMA and HIMA [33, 36, 123-131], so the specific contribution of either type remains uncertain.

The present findings provide several hypotheses that warrant investigation in future rehabilitation studies. Given the longer time-to-task failure observed during PIMA in appendicular muscles, this mode may be advantageous in situations where prolonged force production or sustained muscle activation is desirable. Conversely, the greater sensorimotor demands associated with HIMA may be beneficial in rehabilitation contexts that emphasize postural control, dynamic stability, or neuromuscular coordination, particularly in neurological populations.

Bauer et al. [12] proposed differentiating between the two isometric types according to patient needs; whether this improves patient outcomes warrants investigation. The findings regarding maximal holding capacity, together with the injury prevention considerations above, raise the hypothesis that rehabilitation might benefit from emphasizing muscle stability and adaptation to external loads rather than maximal strength alone. However, direct comparative studies are needed before specific recommendations can be made.

#### Sports performance

Isometric training for sports performance has gained recent popularity as more evidence on its ability to improve various sports performance indicators has become available [1, 34, 35, 132, 133]. However, most of these intervention studies used PIMA [132-135], with only a limited number employing HIMA [34, 35]. Additionally, no study has directly compared the longitudinal effects of PIMA- and HIMA-based training on sports performance outcomes. Hence, it may be challenging for practitioners to determine which method to use at various training cycle phases. Nevertheless, the acute physiological differences identified in the present review provide a framework for generating several testable hypotheses regarding training prescription. Firstly, HIMA may be preferred during general preparation, where the training objective is often to increase the overall strength required to endure the demands of subsequent phases, and to reduce the risk of injury as training intensity increases. This suggestion is based on the greater increase in synergistic muscle activation during HIMA [4, 63, 77, 79], which is vital in maintaining joint integrity during intensive movements [22]. Furthermore, appendicular muscles can sustain PIMA longer than HIMA at a given intensity, while the opposite finding was true for the trunk muscles. During the general preparation phase, coaches plan for higher training volumes, meaning that muscle action duration would be longer if isometric training is included. If PIMA were used during an exercise such as the isometric squat, the adaptation of the lower limb may be limited by the duration for which the trunk muscles could sustain a given force. However, if HIMA were to be used, the trunk muscles would likely be able to maintain the given force for as long as, or longer than, the lower limb. Hence, the adaptation to the lower limb would be less limited by the trunk. Secondly, during the training phase, PIMA would likely be preferred, as it involves exerting force against an immovable object, allowing an athlete to exert maximal force rapidly. Similarly, greater maximal torques during PIMA than HIMA were reported [21, 22, 25]. However, whether these acute differences translate into superior long-term training adaptations remains unknown.

While there is rarely only one mode of muscle action in sports, brief or prolonged isometric action is typical in many sports. Hence, the decision to use PIMA or HIMA in training may also depend on the specific sporting action(s) to be prepared for. For example, sports actions under high external loads with relatively static conditions, such as a rugby scrum or a grappling/wrestling maneuver, may be best trained using PIMA. Likewise, actions that require holdings, such as the static position held during a springboard dive or various gymnastic maneuvers, may be best prepared for using HIMA. Finally, if improving rate of force development is paramount, both PIMA and HIMA with rapid muscle actions can be utilized [136]. While it is relatively simple to perform PIMA with rapid intent, practitioners may lack insight into how to conduct rapid HIMA, including rapid landings or unconventional exercises (e.g., rapid decelerations, hamstring ‘switches’, oscillatory training), which are currently lacking in scientific literature. Furthermore, while several examples of multi-joint HIMA exist (**Online Resource 8**) [34, 35], no longitudinal studies have compared PIMA and HIMA via closed-chain movements, presenting an exciting research opportunity.

### Limitations and future research directions

While the primary aims were accomplished, there are several limitations. First, we included only studies that directly compared PIMA and HIMA. While it would have been exceptionally time-consuming, all studies employing PIMA or HIMA could have been identified and synthesized. While enough studies utilized TTF as a dependent variable for confident meta-analyses, far fewer studies reported HR, MAP, or RPE. Therefore, readers should be aware of potential limitations. Similarly, the between-study variability in neurological (e.g., EEG, TMS) and neuromuscular (e.g., sEMG) assessment methodology prevented us from performing further meta-analyses. Moreover, the small number of studies focusing on the same objective (e.g., brain activity, tendon oscillations, max torque) makes authoritative statements difficult. Other methods may uncover further insights, including fMRI, fascicle dynamics, echo intensity, shear wave elastography, tendon creep, and tensiomyography. The ‘stability-of-position’ factor could not be accounted for in most studies (i.e., HIMA is typically more unstable than PIMA); only eight studies used setups with similar stability between tasks. Thus, aligning the position stability during both functions would be advisable. Readers should also be aware that the experimental pain trials included in this review were conducted in highly controlled laboratory settings, which may not be directly applicable to clinical settings.

The available evidence is clustered within a small number of research groups: 16 of the 28 appendicular TTF outcomes originate from a single laboratory, the mechanistic outcomes (e.g., EEG and coherence measures) are similarly concentrated, and the comparisons of maximal torques of HIMA (HIMA_max_) derive entirely from members of the present authorship team. This concentration reflects the emerging nature of the field, but it raises the possibility that study design, terminology, and interpretation are shaped by shared theoretical perspectives. Sensitivity analyses indicated that the direction and statistical significance of the appendicular TTF effect were retained after exclusion of any single study, first author or laboratory, and of the two collaborating laboratories jointly; omitting the outcomes of the authors’ own laboratory did not change the estimate (**Online Resource 7**). The magnitude of the appendicular effect, however, was larger within the largest laboratory, which may reflect methodological composition, particularly the predominance of low loading intensities, as much as investigator effects. For overall TTF, including the two axial-muscle outcomes, the estimate after joint exclusion of the two collaborating laboratories was no longer statistically significant (*g* = −0.27, 95% CI −0.67 to 0.14; **Online Resource 7**). Independent replication using standardized protocols therefore remains a priority.

Risk-of-bias was assessed using the revised JBI tool. Within-study reliability of the outcome measures was frequently not reported (Q7); this indicates uncertainty about measurement error, including whether it affected both conditions equally. Our appraisal did not comprehensively assess allocation of task order, period or carryover effects, selective reporting or investigator allegiance. Within-participant comparisons account for stable differences between individuals, but do not eliminate fatigue, learning or incomplete recovery as explanations for task differences. Furthermore, counterbalancing without explicitly reported random allocation does not establish a randomized sequence, and fixed-order and historical comparisons provide less protection against confounding. Together with the common high starting point, an explicit adaptation of GRADE, the limitations of our risk-of-bias assessment may have resulted in overly optimistic certainty ratings. The funnel plot was inspected visually, but formal statistical tests of asymmetry were not performed because heterogeneity, dependent effects, and the relationship between standardized effects and their standard errors complicate interpretation. Estimates with large standard errors were predominantly subgroup cells from larger studies, and null findings were reported; nevertheless, selective reporting or publication bias cannot be excluded.

Many primary studies did not report effect sizes, requiring calculation from summary data or figure digitization, which introduces additional uncertainty. Within-participant correlations were back-calculated or assumed where not reported; the pooled estimates were robust to the assumed correlation, although the statistical significance of single HR and MAP differences depended on it (**Online Resource 6**). These analyses assess sensitivity to the variance assumptions, not to allocation, carryover or other sources of bias. The certainty ratings apply to the laboratory outcomes assessed and do not establish clinical benefits or long-term training adaptations.

The mean intensity of the included studies was far lower than nearly all but the most conservative rehabilitation or sports performance guidelines. While these low loads allowed for greater durations and, thus, more time intervals for comparison, future studies should examine more practically relevant intensities. An even more explicit limitation is the complete absence of studies comparing PIMA and HIMA during multi-joint movements (e.g., squat, leg press, pull-up, bench press). Researchers could also compare the two types of isometric muscle action over multiple days to examine the time course of fatigue and recovery. Moreover, investigations on how external and internal stimuli might influence HIMA and PIMA could be relevant for gathering further information on underlying control mechanisms and for diagnostics, injury mitigation and rehabilitation. Most importantly, longitudinal, randomized controlled trials are required to ascertain if PIMA or HIMA have any measurable advantage over the other for use in diagnostics or for inducing various rehabilitation or sports performance adaptations.

## Conclusions

This systematic review and meta-analysis advances the understanding of isometric muscle actions by delineating two distinct types—PIMA and HIMA—and synthesizing evidence that they differ in neural, biomechanical, and functional ways. The clearest evidence, supported by meta-analysis with moderate certainty, is a longer TTF for PIMA than HIMA in appendicular muscles at the same absolute intensity, whereas certainty was low for overall TTF and for the opposite axial-muscle finding. Most other task-dependent differences were supported by evidence of low to very low certainty. The neurophysiological findings, largely based on single or few studies, are consistent with the hypothesis that HIMA involves more complex neural and neuromuscular control than PIMA, possibly resembling the processing of lengthening and shortening actions, respectively (‘stopped-action’ hypothesis); this evidence is, however, of very low certainty and requires direct testing. Faster increases in selected neuromuscular and cardiovascular responses during HIMA, alongside heightened sensorimotor modulation, also motivate hypotheses concerning diagnostics, injury prevention and rehabilitation, particularly under increasing external loads.

When integrated with broader physiological, biomechanical, and psychological research, the acute differences generate testable hypotheses for training and rehabilitation: that HIMA may offer a time-efficient means of inducing musculoskeletal, neural, and cardiovascular adaptations, and that PIMA may be more effective for sustained activation and promoting agonist-specific neuromuscular adaptations owing to its greater force capacity. Whether these acute differences translate into distinct long-term adaptations is unknown, as no study has compared PIMA- and HIMA-based training longitudinally.

In addition to the quantitative syntheses, the scoping component of this review identified several important gaps in the literature, including the absence of multi-joint investigations, the limited number of longitudinal intervention studies, the wide methodological heterogeneity in neurophysiological assessments, and the paucity of studies employing advanced imaging or biomechanical techniques. These findings complement the meta-analyses by providing a broader overview of the current evidence base, underscore the need for consistent protocols and improved reporting, and help to prioritize future research directions.

Collectively, the findings challenge the traditional view of isometric muscle actions as a unitary construct and highlight potentially important implications for sports medicine and rehabilitation, where isometrics are widely used to enhance recovery, prevent injury, and improve performance. Incorporating both HIMA and PIMA into assessment, training, and rehabilitation might enable practitioners to target complementary adaptations, individualize care, and develop goal-specific interventions; as the acute laboratory differences support hypotheses rather than recommendations, this should be tested in future randomized trials in clinical and sport contexts. We suggest that the distinction between PIMA and HIMA, together with the limitations of the evidence, be incorporated into research and education, while its implications for clinical practice undergo further evaluation.

## Data Availability

All data produced in the present study are available upon reasonable request to the authors

## List of abbreviations

AF: Adaptive Force
AUC: Area under the curve
CV: Coefficient of variation
EEG: Electroencephalography
EMG: Electromyography
aEMG: Electromyographic amplitude
sEMG: Surface electromyography
GRADE: Grading of Recommendations Assessment, Development, and Evaluation
HIMA: Holding isometric muscle actions
HR: Heart rate
ISI: Interspike interval
JBI: Joanna Briggs Institute
MAP: Mean arterial pressure
MEP: Motor evoked potential
MMG: Mechanomyography
MTG: Mechanotendography
MVIC: Maximal voluntary isometric contraction
PIMA: Pushing/pulling isometric muscle actions
PRISMA: Preferred Reporting Items for Systematic Reviews and Meta-Analyses
REML: Restricted maximum likelihood
RMS: Root-mean-squared
RPE: Ratings of perceived exertion
SD: Standard deviation
SE: Standard error
SWiM: Synthesis Without Meta-analysis
TMS: Transcranial magnetic stimulation
TTF: Time to task failure

## Declarations

### Ethics approval and consent to participate

Not applicable

### Availability of data and materials

The datasets used and/or analyzed during the current study are available from the corresponding author on reasonable request.

### Authors’ contributions

DJO conceptualized the review topic and formed the research team. DJO and ARN fine-tuned the research question and review methods, performed the literature search, and conducted initial screenings. DJO, DL, LVS, AON and ARN assessed studies for inclusion. DJO and ARN conducted the revised JBI appraisal, with DL adjudicating disagreements. DJO and ARN performed the GRADE assessment. DJO, ARN, DL and LVS extracted data, and FNB checked the extracted data for accuracy. DJO, LVS and ARN performed the statistical analyses and created the figures; LVS performed the sensitivity and robustness analyses; DJO, ARN, DL, and AON completed the original risk-of-bias appraisal; DJO and ARN completed the revised risk-of-bias appraisal, and GRADE assessment. DJO wrote the first draft of the title, abstract, introduction and methods. DJO and LVS wrote the first draft of the results. All authors contributed to the discussion’s first draft, critically revised the manuscript, and approved the final version.

## Acknowledgments

Not applicable

## Online Resources

– **Online Resource 1** Risk-of-bias appraisal of the included studies using the revised JBI critical appraisal tool for quasi-experimental studies: operational definitions, decision rules, and study-, outcome-, and result-level judgments.
– **Online Resource 2** Evidence profile (GRADE) for the meta-analyzed outcomes and the narratively synthesized outcome groups comparing PIMA and HIMA.
– **Online Resource 3** Summary of studies comparing performance and biomechanical parameters between pushing (PIMA) and holding (HIMA) isometric muscle actions.
– **Online Resource 4** Summary of studies comparing neural and neuromuscular parameters between pushing (PIMA) and holding (HIMA) isometric muscle actions.
– **Online Resource 5** Summary of studies comparing cardiovascular and metabolic parameters between pushing (PIMA) and holding (HIMA) isometric muscle actions.
– **Online Resource 6** Sensitivity analysis of the pooled effects for different assumed within-participant correlations (*r*) between HIMA and PIMA.
– **Online Resource 7** Robustness analyses for time to task failure (TTF).
– **Online Resource 8** Examples of multi-joint pushing (PIMA; left) and holding (HIMA; right) isometric muscle actions: back squat, hip extension, and bench press. In the pushing variants, force is exerted against an immovable resistance (barbell fixed to the rack); in the holding variants, the free barbell load is held in a set position.

